# The effectiveness of non-pharmacological treatments for auditory verbal hallucinations in schizophrenia spectrum disorders: a systematic review and meta-analysis

**DOI:** 10.1101/2025.05.08.25327225

**Authors:** Melis Cobandag, Natasha Sigala

## Abstract

**Background:** Schizophrenia is a chronic severe mental illness affecting 24-million people globally, associated with a life expectancy 15 years shorter than the general population. Approximately 70% of people with schizophrenia experience auditory verbal hallucinations (AVHs), i.e. ‘hearing voices’. Current treatment approaches remain unsuccessful at treating AVHs in up to 30% of cases.

**Aims:** This systematic review and meta-analysis evaluated randomised controlled trials (RCTs) of potential non-pharmacological treatments for AVHs in schizophrenia spectrum disorders. The effectiveness of emerging treatments was assessed and gaps in research were identified, with implications for clinical practice.

**Methods:** A literature search was performed between 2013-2024 across five databases: PubMed, Embase, PsycINFO, Medline, and Web of Science. The meta-analysis included 45 studies, based on predefined criteria and an assessment for bias. Effect sizes (Hedge’s g) were calculated for each study and overall, using a random effects model with 95% confidence intervals. The study was conducted in accordance with PRISMA guidelines and was pre-registered (PROSPERO ID: CRD42024598615).

**Results:** Our sample included 2,314 patients and fourteen different interventions. The calculated overall mean effect size for all interventions was −0.298 (95% CI, [−0.470, −0.126]), representing a medium size and statistically significant effect. Subgroup analyses revealed that both AVATAR therapy and cognitive behavioural therapy (CBT) subgroups had medium size and statistically significant effects. Conversely, repetitive transcranial magnetic stimulation (rTMS) and transcranial direct current stimulation (tDCS) showed small size and not statistically significant effects.

**Conclusions:** AVATAR therapy has the strongest evidence for treating AVHs, highlighting the need for large-scale RCTs and integration into treatment guidelines. CBT requires standardisation in methodology for reliability, and clinical guidelines should focus on symptomology rather than diagnosis. Acceptance and commitment therapy shows promise but requires further high-quality RCTs. Non-invasive brain stimulation techniques require further trials before they can be considered for clinical use.

## Introduction

Schizophrenia is a chronic and severe mental illness affecting 24-million people globally,^1^ accounting for approximately 1% of the population.^2^ On average, people with schizophrenia have a life expectancy 15 years shorter than the general population, and a risk of death by suicide of up to 10%.^3^ Although symptoms and their severity vary widely between individuals, contributing to the complexities of treatment, one frequently experienced symptom is auditory verbal hallucinations (AVHs), i.e. ‘hearing voices’, present in approximately 70% of people with schizophrenia.^4^ AVHs are a key contributor to schizophrenia-related morbidity and mortality by reducing quality of life and elevating the risk of violence and suicide.^5, 6^ Therefore, managing AVHs is critical to reduce the morbidity of this illness, with implications for potential uses in other conditions in which AVHs are a prominent symptom, such as in schizoaffective disorder, or bipolar affective disorder.^7^

The current first-line treatment for schizophrenia is antipsychotic medication,^8^ however for 30% of patients these AVHs persist despite treatment.^9–11^ People with schizophrenia can often lack insight into their condition, impacting medication adherence,^12^ which is compounded by the potential for severe adverse effects of antipsychotics. Some serious side effects include tardive dyskinesia,^13^ prolonged QT syndrome,^14^ and agranulocytosis which is of particular concern when using clozapine, an antipsychotic reserved for treatment resistant schizophrenia.^15^

The limited efficacy and serious side effects of antipsychotics in treating AVHs underscores a need for alternative or additional treatments, prompting a recent shift towards non-pharmacological interventions. Examples of these interventions include cognitive behavioural therapy (CBT),^16^ a talking therapy that encourages change in cognition and behaviour, and repetitive transcranial magnetic stimulation (rTMS), a non-invasive brain stimulation technique that may modulate the neural activity implicated in AVHs.^17^ Non-pharmacological treatments are often trialled alongside antipsychotic medications.

However, there is still uncertainty surrounding the efficacy of non-pharmacological treatments, in addition to a lack of specifically tailored guidance for their implementation in clinical settings. Reasons for this may include conflicting evidence across various studies, summarised in previous systematic reviews focusing on specific types of non-pharmacological treatments.^18–21^ Moreover, previous systematic reviews have focused on specific subtypes of non-pharmacological interventions for AVHs, lacking in critical evaluation and comparison between different types of non-pharmacological interventions.^18–21^ Additionally, such studies have not exclusively reviewed randomised controlled trials (RCTs),^18^ which are the most methodologically robust way to test an intervention.^22^ Therefore, this study seeks to provide an evaluation of recent RCTs testing non-pharmacological treatments for AVHs in SSDs. To the best of our knowledge, this is the first meta-analysis to be conducted on all non-pharmacological interventions for AVHs in SSDs.

### Aims

The specific objectives of this systematic review and meta-analysis were:

1. To evaluate the overall efficacy of non-pharmacological interventions for AVHs based on RCTs conducted in the last decade.
2. To compare the effectiveness of different types of interventions, such as CBT, rTMS and AVATAR therapy, measured by validated rating scales.
3. To assess the methodological quality and limitations of included studies.
4. To identify gaps in research and provide recommendations for future studies and clinical guidelines.

## Methods

### Search strategy, selection criteria and quality assessment

This systematic review and meta-analysis was performed in accordance with the Preferred reporting Items for Systematic Reviews and Meta-Analyses (PRISMA) guidelines^23^ (see supplementary Figure S1). The protocol was preregistered on PROSPERO (ID: CRD42024598615) on Nov 18^th^ 2024.

Five electronic databases were searched on the 25^th^ of November 2024. Searches were performed using MeSH terms and Boolean operators, outlined in Table 1. Full search strings are provided in supplementary Table S2. Additional search filters were applied, detailed in Table 2.

**Table 1.**
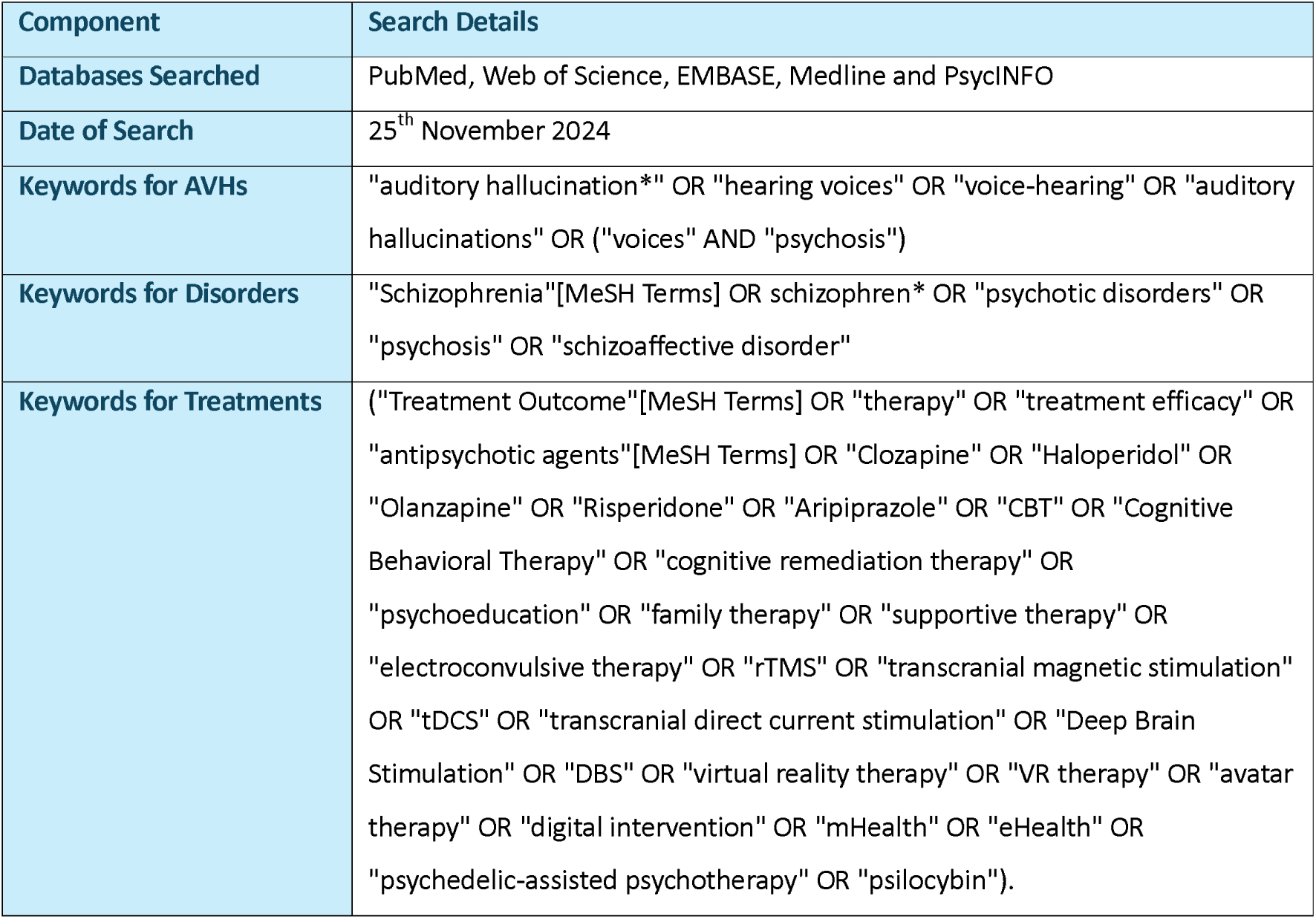
Search details.

| Component | Search Details |
| --- | --- |
| Databases Searched | PubMed, Web of Science, EMBASE, Medline and PsycINFO |
| Date of Search | 25<sup>th</sup> November 2024 |
| Keywords for AVHs | "auditory hallucination*" OR "hearing voices" OR "voice-hearing" OR "auditory hallucinations" OR ("voices" AND "psychosis") |
| Keywords for Disorders | "Schizophrenia"[MeSH Terms] OR schizophren* OR "psychotic disorders" OR "psychosis" OR "schizoaffective disorder" |
| Keywords for Treatments | ("Treatment Outcome"[MeSH Terms] OR "therapy" OR "treatment efficacy" OR "antipsychotic agents"[MeSH Terms] OR "Clozapine" OR "Haloperidol" OR "Olanzapine" OR "Risperidone" OR "Aripiprazole" OR "CBT" OR "Cognitive Behavioral Therapy" OR "cognitive remediation therapy" OR "psychoeducation" OR "family therapy" OR "supportive therapy" OR "electroconvulsive therapy" OR "rTMS" OR "transcranial magnetic stimulation" OR "tDCS" OR "transcranial direct current stimulation" OR "Deep Brain Stimulation" OR "DBS" OR "virtual reality therapy" OR "VR therapy" OR "avatar therapy" OR "digital intervention" OR "mHealth" OR "eHealth" OR "psychedelic-assisted psychotherapy" OR "psilocybin"). |

**Table 2.** Search filters applied to databases, and number of studies yielded.

| Database | No. initial search results | Additional filters | No. of results with additional filters |
| --- | --- | --- | --- |
| PubMed | 930 | Randomised controlled trial<br>Human<br>English | 150 |
| Web Of Science | 1228 | English<br>Add "controlled trial" to search in all fields<br>Publication type: Article | 208 |
| PsycINFO | 1539 | Clinical trial<br>English<br>Peer reviewed<br>Adult | 75 |
| Medline | 1249 | Humans<br>English<br>Randomised controlled trial | 138 |
| EMBASE | 4811 | Human<br>English language<br>Randomised controlled trial<br>Publication type: Article<br>Source type: Journal | 146 |
| **Total:** | 9757 |  | 717 |

Inclusion criteria for studies were a) Adults aged ≥18 years, b) majority of participants with a formal diagnosis of schizophrenia or other psychotic disorder, c) RCTs, d) any treatment/therapy for auditory hallucinations, e) papers published in English. Papers including participants with substance misuse or a coexisting neurological condition were excluded. All other study types that were not RCTs were excluded.

The Standard Quality Assessment Criteria for Evaluating Primary Research Papers from a Variety of Fields, also known as the QualSyst Tool^24^ (detailed in supplementary Table S3), was used to evaluate the risk of bias of each article which met the inclusion criteria. Risk of bias scores for each study were calculated by both reviewers independently, then compared to evaluate inter-operator agreement, and an inclusion threshold of 0.55 was used. If an article met all eligibility criteria but lacked data on the AVH measure, corresponding authors were contacted. Out of fourteen corresponding authors contacted, two responded with the post-treatment data required.^25, 26^ The other eleven studies were not included in this meta-analysis.^27–37^ Another paper was excluded^38^ due to the author responding to explain that their paper reanalysed the data reported in study 31. QualSyst bias assessment scores for each study are provided in supplementary Table S4.

### Data synthesis and analysis

Data extraction started on the 17th of December 2024. The following information was extracted from each article: setting, patient sex, age, ethnicity, diagnosis, other treatments participants received (other than the intervention of interest), intervention tested, control condition, frequency and dosage of intervention, AVH scoring system, number of participants in each group, mean and standard deviations of the AVH score at baseline and post-treatment, and a summary of key findings.

The primary outcome measure was the difference in post-treatment AVH scores between the control group and intervention group, where the AVH score measured at least one of the following: frequency, distress or intensity of AVH. The data required to calculate change scores and standard deviations were inconsistently reported; therefore, to avoid making assumptions, using post-treatment AVH scores was most suitable. This is appropriate in the absence of change score data, given that there are minimal baseline differences between the groups. Baseline scores were carefully considered for all studies, and were similar across groups, providing a suitable and comparable measure of treatment effect.

Analyses were performed using the software Comprehensive Meta-Analysis Version 4 (CMA V4),^39^ which assessed the heterogeneity of the studies, and calculated the effect size for each study and overall. Forrest plots were produced, using a random effects model with 95% confidence intervals. The heterogeneity of the studies (the outcome variability due to clinical and methodological differences) was measured with Higgins’s I^2^ statistic.^40^ Higgins’s I^2^ represents the percentage of variation between the sample estimates that is due to heterogeneity rather than a sampling error.^41^ Subgroup analyses was performed for an intervention reported in at least 5 studies.

Risk of publication bias was independently assessed by both reviewers, through visual inspection of the funnel plot, which was created using CMA V4.

## Results

The initial search produced 717 records. After removal of duplicates, titles and abstracts of 399 articles were screened. If titles and abstracts appeared relevant, full texts were retrieved. 77 full-text articles were assessed for eligibility. 45 articles met all eligibility criteria and were included in the meta-analysis (Fig.1).

**Fig. 1.**
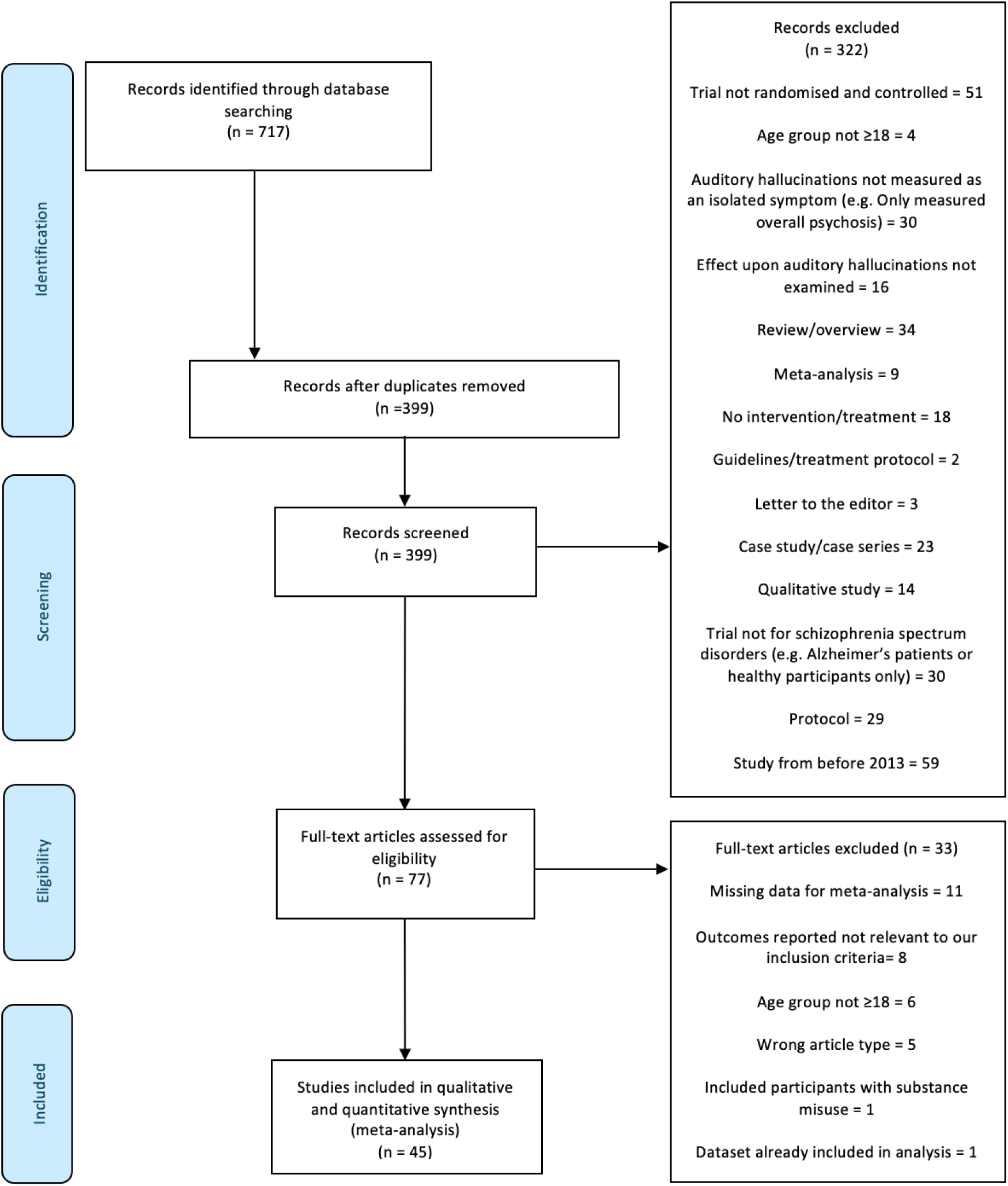
PRISMA flow diagram for study inclusion.

**Fig. 2.**
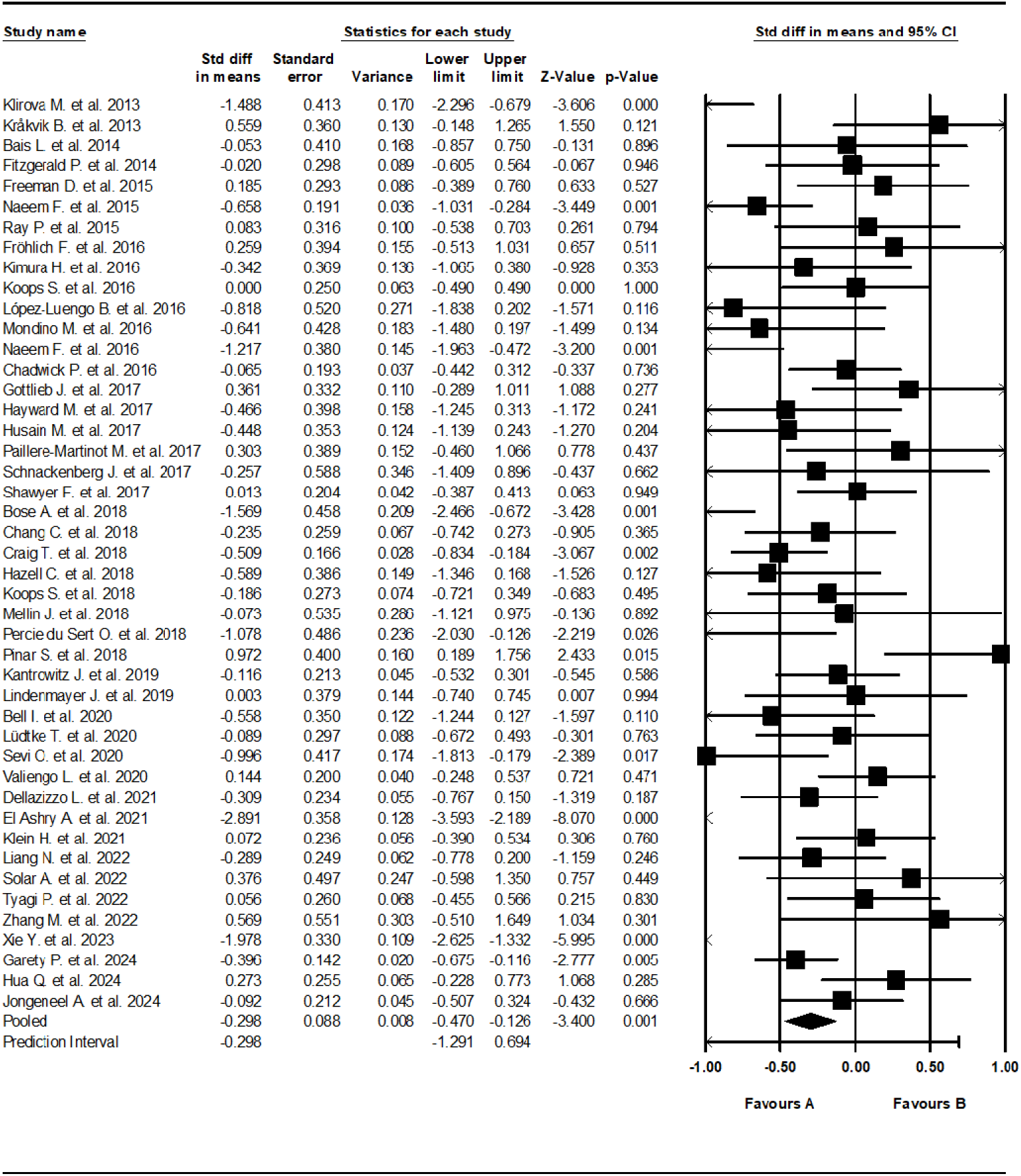
Forest plot of standardized mean differences (SMD) between post-intervention AVH scores in those who received an intervention for AVHs (A), versus the control group (B). Negative effect sizes indicate lower (improved) AVH scores in the intervention group compared to the control group.

### Study characteristics and participant characteristics

Studies identified included the following 14 types of treatment: rTMS, CBT, transcranial direct current stimulation (tDCS), AVATAR therapy, transcranial alternating current stimulation (tACS), smartphone app treatments, acceptance and commitment therapy (ACT), psychological online intervention (POI), relating therapy (RT), counselling, music therapy (MT), referral to a community psychologist, group mindfulness-based intervention, and rehabilitation. Supplementary Figure S1 reports the number of each intervention type included in the analyses.

For all studies, sample sizes ranged from 12 to 201 participants, with a median of 37. Inclusion criteria across studies involved adults diagnosed with schizophrenia or related disorders, who experienced AVHs. Various AVH scoring systems were used across studies, including the Auditory Hallucinations Rating Scale (AHRS),^42^ Positive and Negative Syndrome Scale – Auditory Hallucinations (PANSS AH), Psychotic Syndrome Rating Scales – Auditory Hallucinations (PSYRATS AH), The Hamilton Program for Schizophrenia Voices Questionnaire (HPSVQ), Delusion and Voices Self-Assessment (DV-SA), and Characteristics of Auditory Hallucination Questionnaire (CAHQ). Each scoring system included a measurement of at least one of frequency, distress or intensity of AVHs. For all AVH scoring systems used, a lower score indicated better AVH outcomes. A change in baseline to post-treatment scores was interpreted as reflecting treatment effects, with a decrease in scores indicating improvement.

Trials were conducted across 20 different countries. Majority of studies (31) were conducted in Western countries, with an emerging representation (12) from Asia. There was an underrepresentation from Africa and Latin America, with only 2 studies conducted in these regions.

In all but two trials,^49^ it was stated that the intervention being explored was administered alongside usual treatments, with a common theme of maintaining stable medication regimes. This approach ensured that any observed changes could be attributed to the intervention, rather than changes in established routine treatments.

Participant demographics and study characteristics are detailed in Table 2 and Table 3 respectively.

**Table 2.**
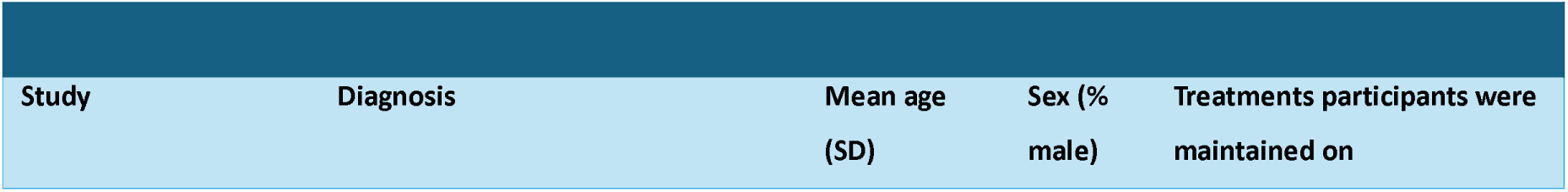
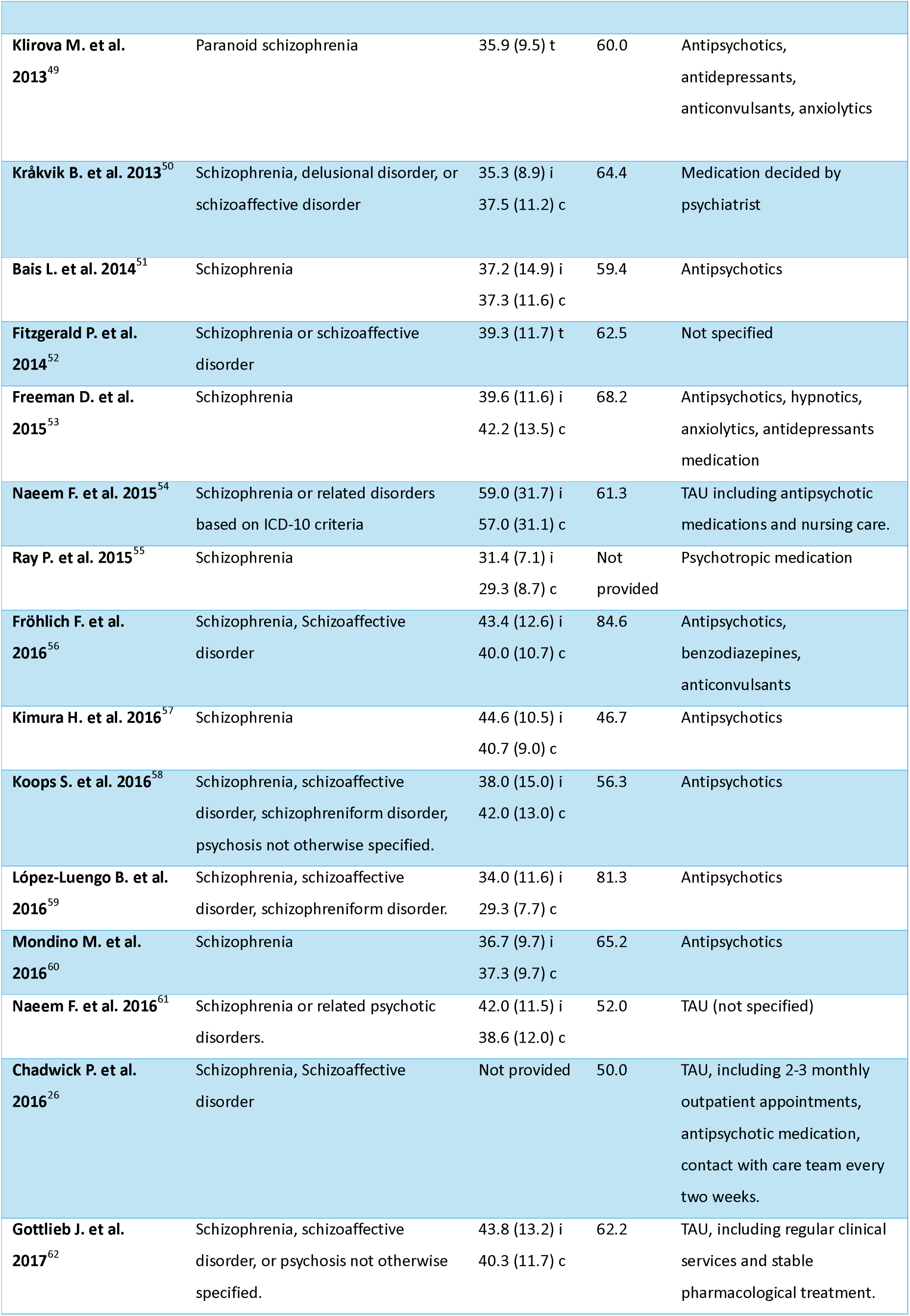
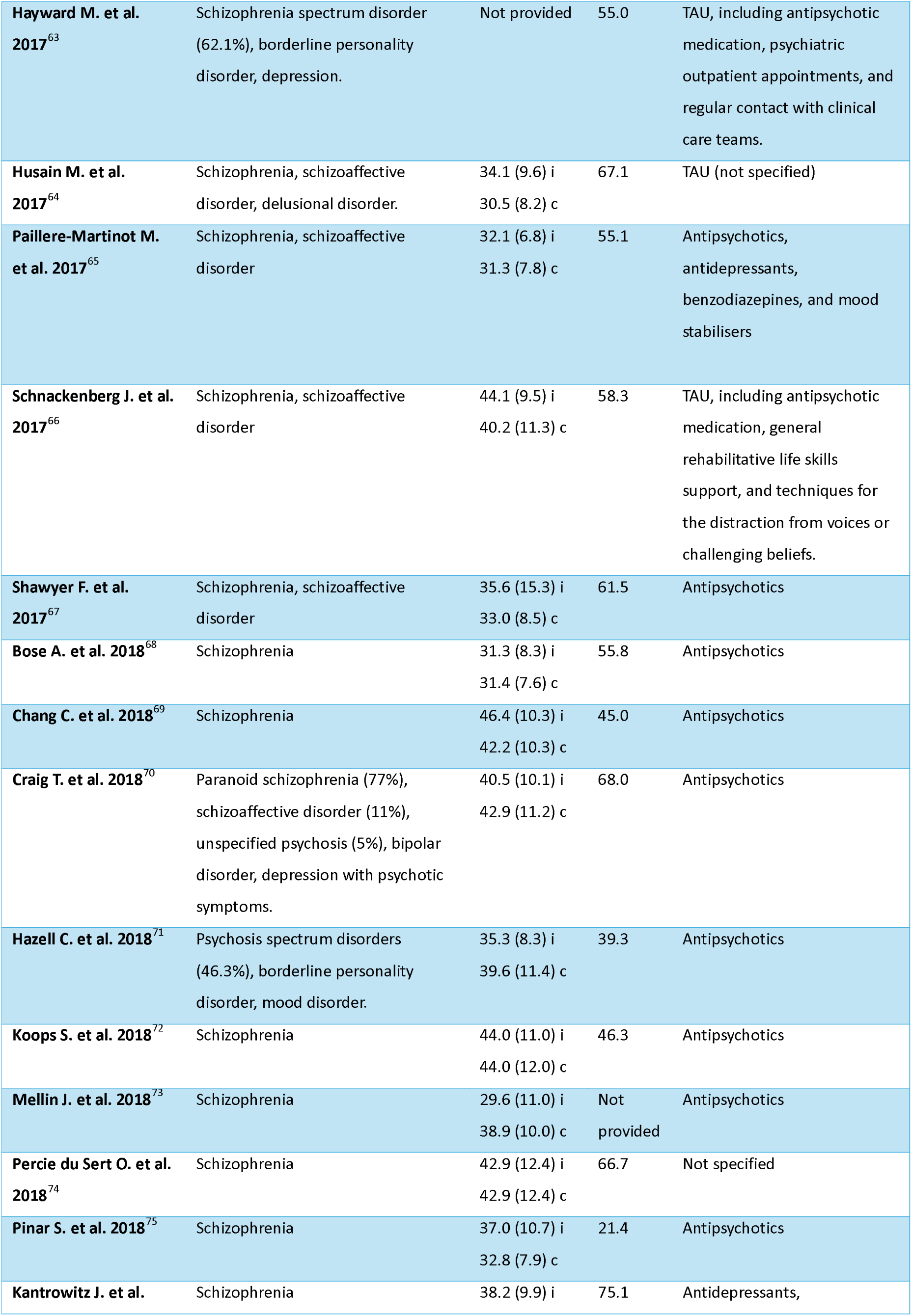
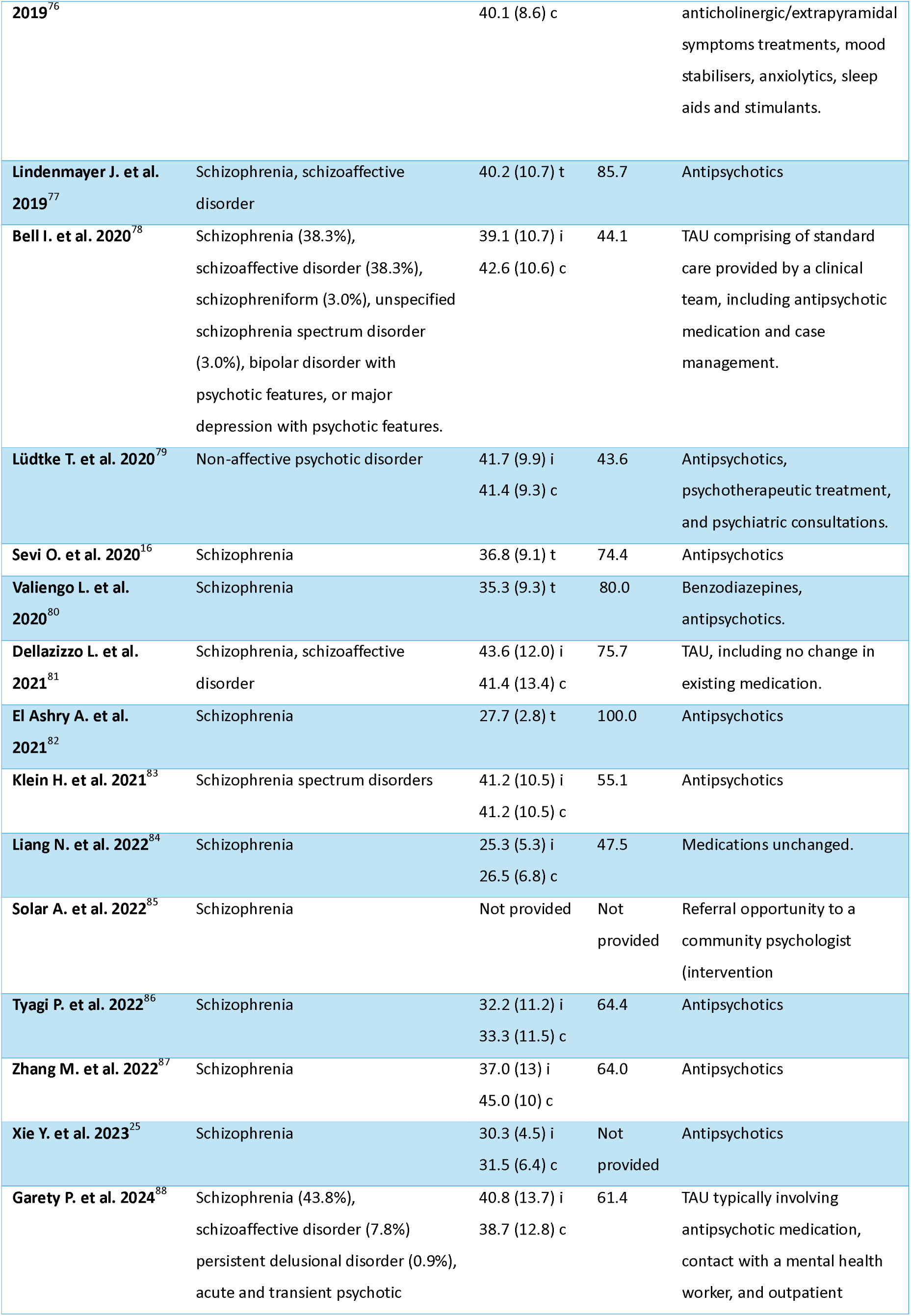
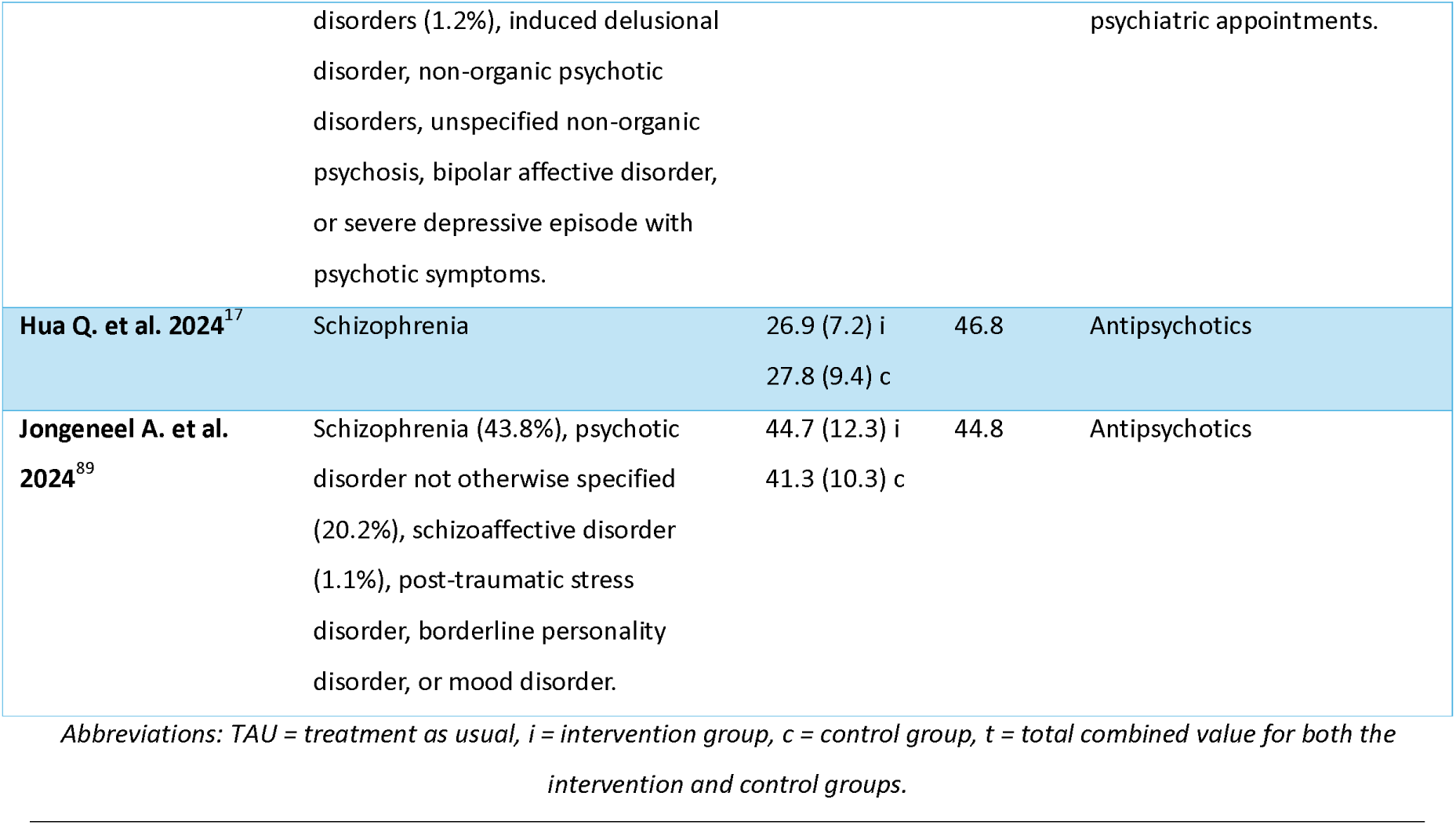
Demographics of included studies. ^16, 17, 25, 26, 49–89^.

**Table 3.**
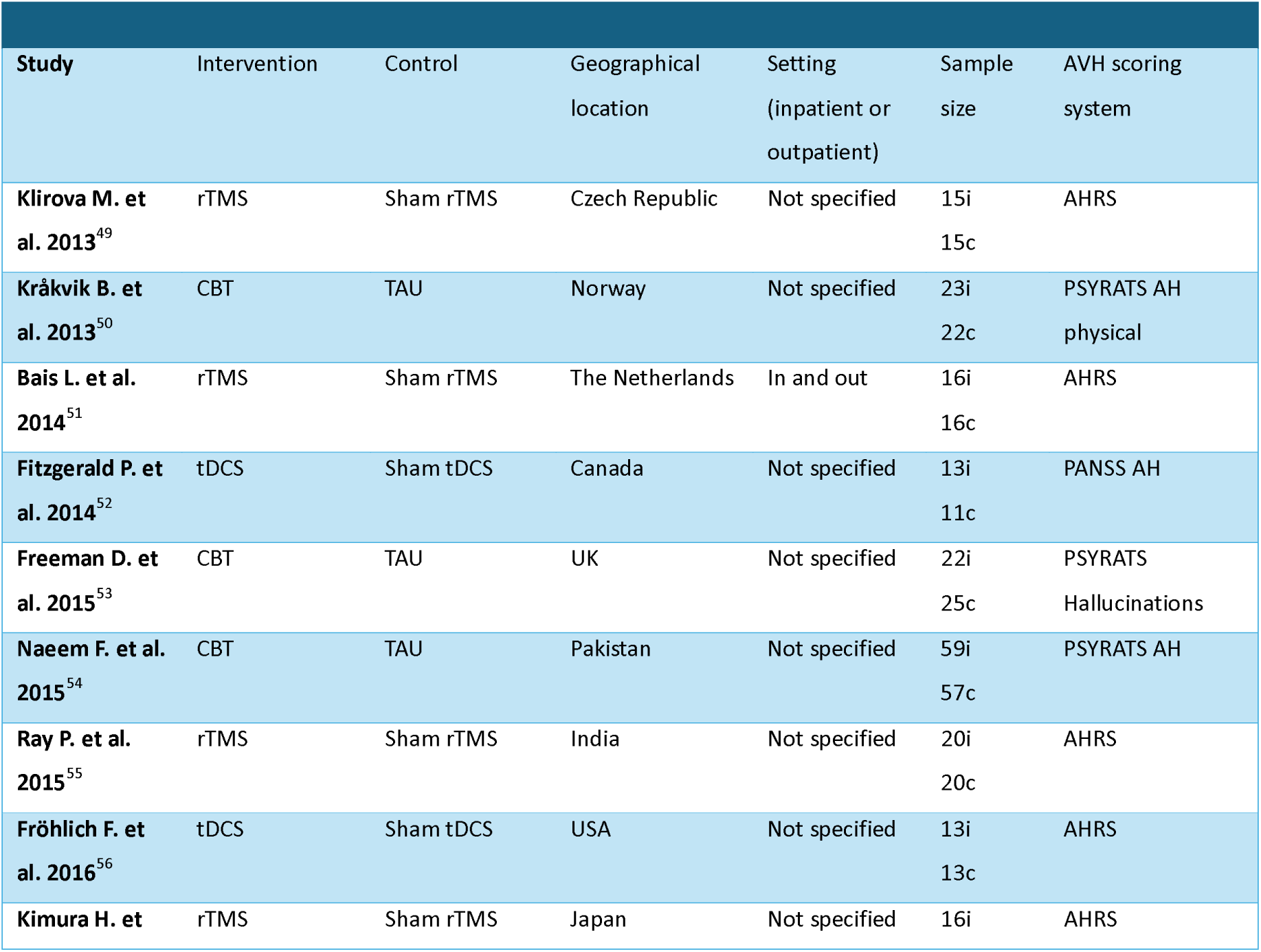
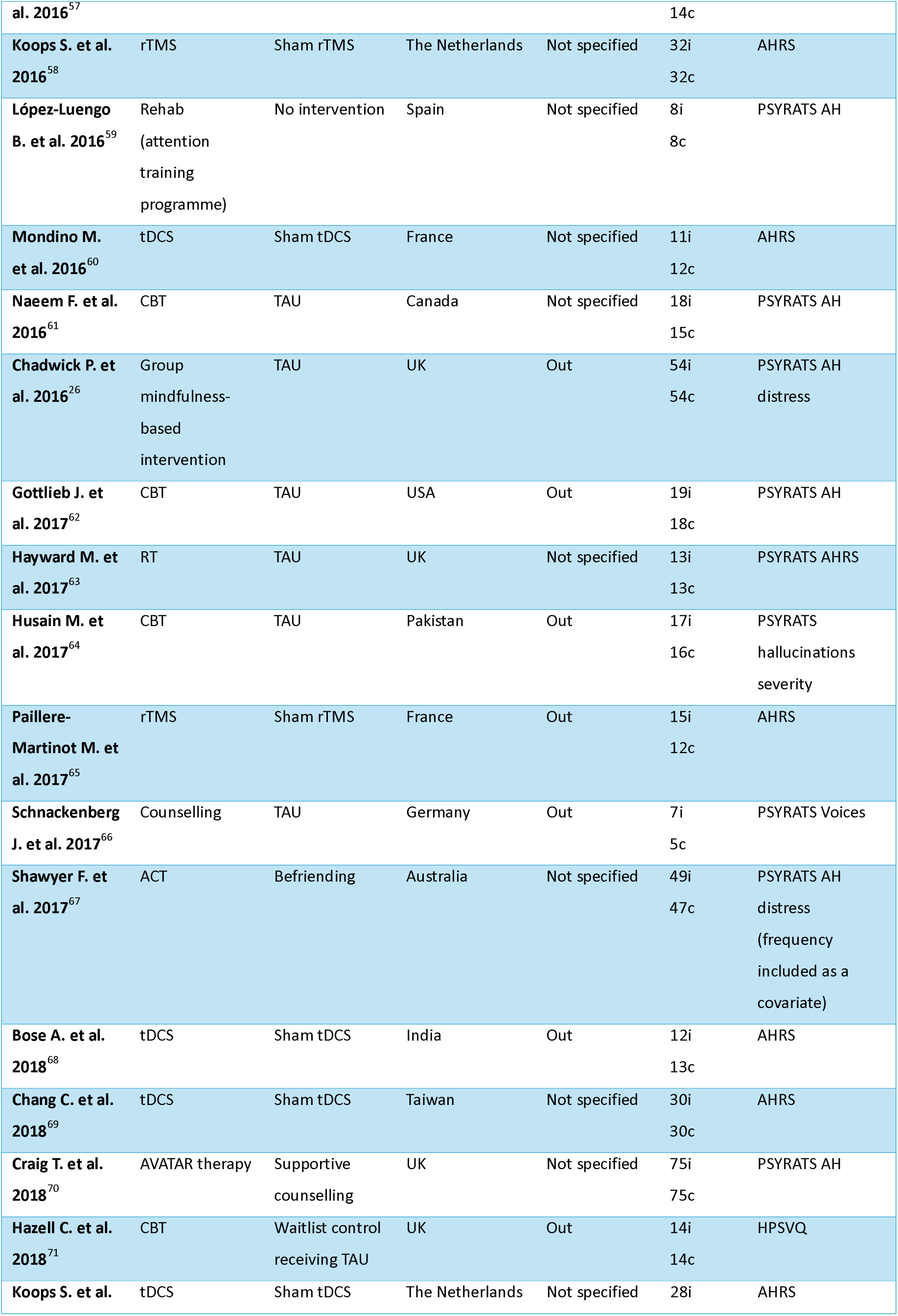
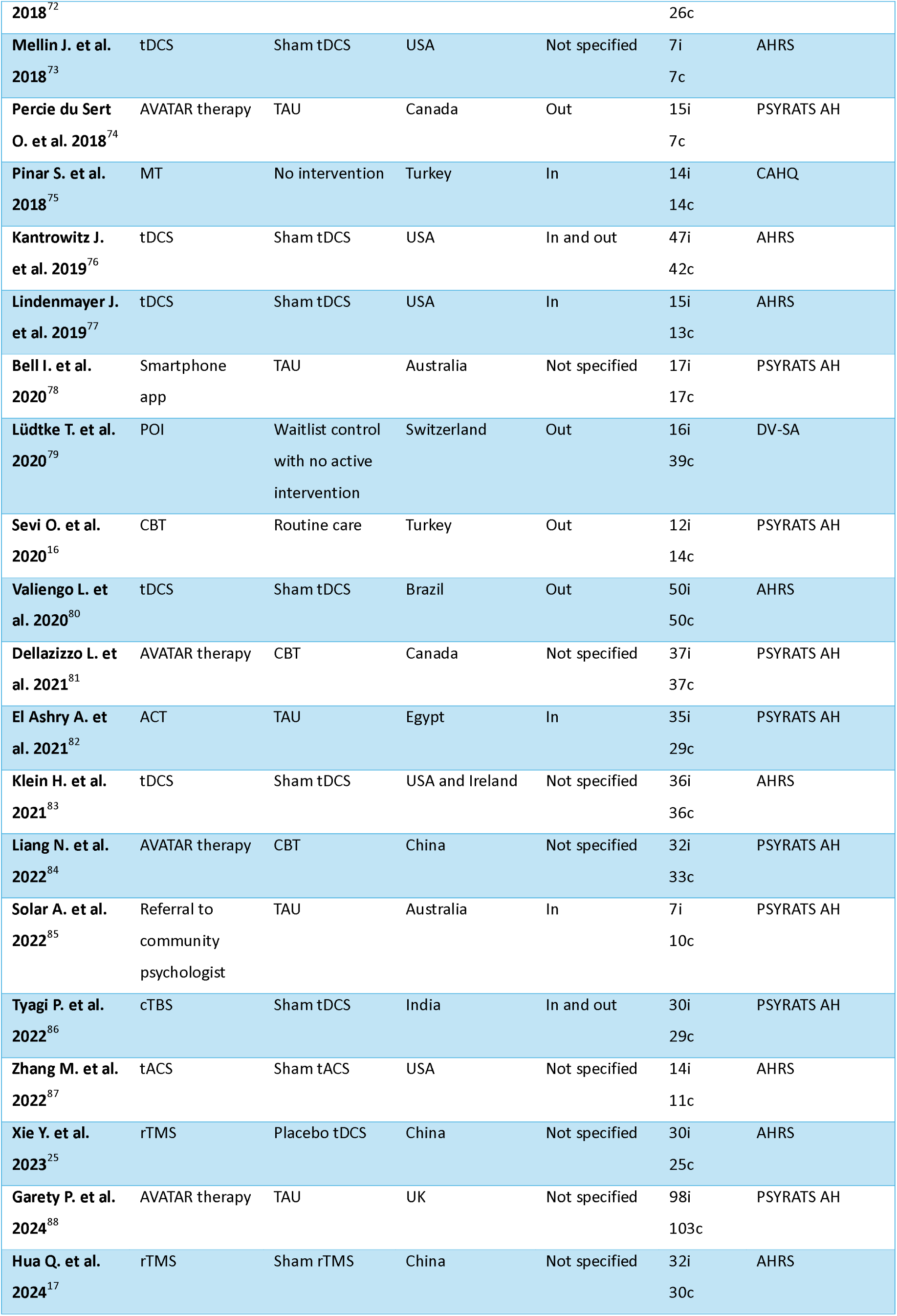
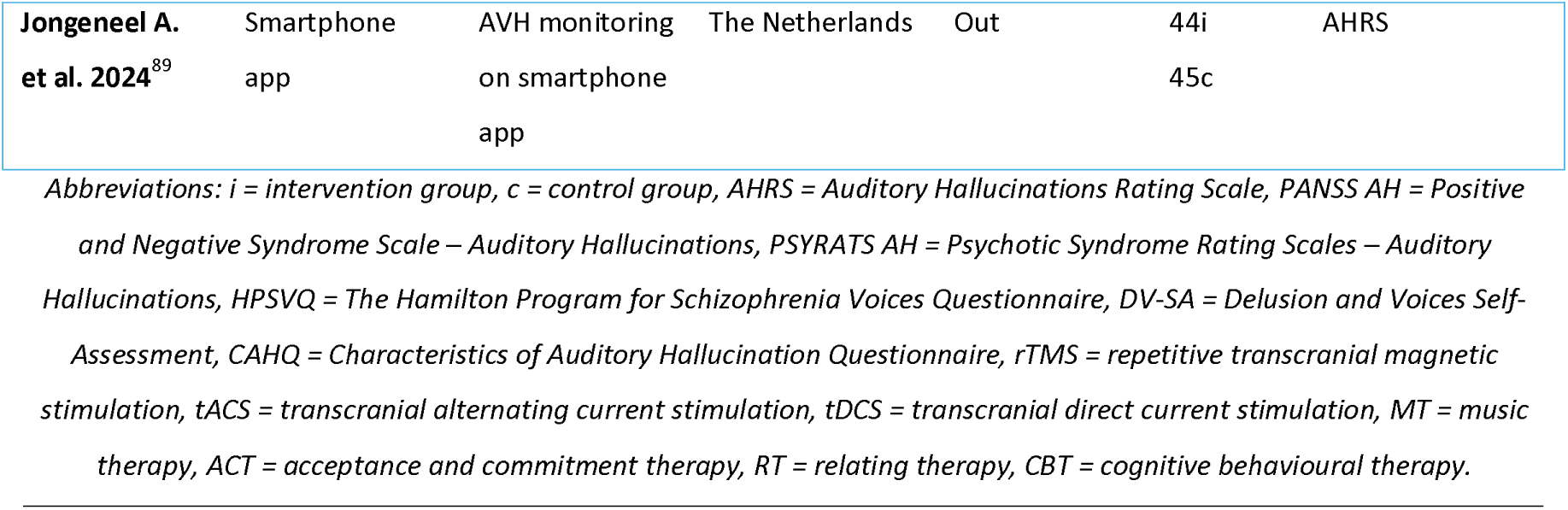
Characteristics of included studies. ^16, 17, 25, 26, 49–89^

### Synthesis of results

#### Overall

Our sample size included 2,314 participants across all studies. The funnel plot (Fig. 3) revealed an even distribution of studies reporting both positive and negative effects, and no bias for studies reporting larger, rather than smaller, effect sizes. An additional precision funnel plot was created to provide a clearer depiction of the relationship between study size and effect size (Fig. S2). Overall, a medium and statistically significant effect size of −0.298 (95% CI, [−0.470, −0.126], Z= −3.400, p= 0.001) was observed across all non-pharmacological interventions. There was significant heterogeneity between studies (Q= 171.566 with 44 degrees of freedom, p < 0.001, I^2^= 74%) indicating substantial variability in effect sizes.

**Fig. 3.**
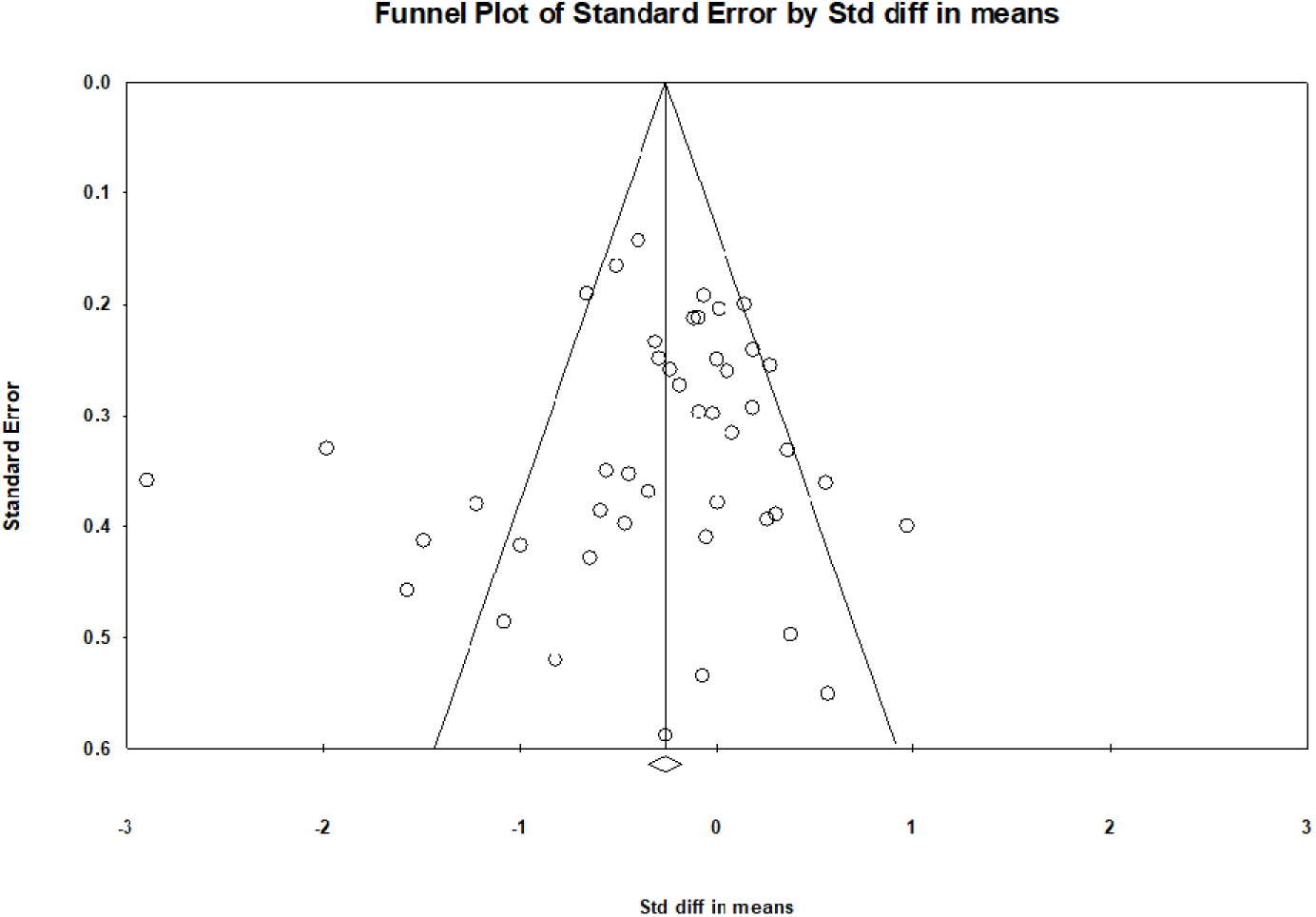
Funnel plot for publication bias.

### Subgroup analysis of interventions

As significant heterogeneity was determined, subgroup analyses of AVATAR therapy, CBT, rTMS and tDCS were performed to explore whether different intervention types influenced effect sizes.

### AVATAR therapy

Five studies tested the use of AVATAR therapy and had sample sizes ranging from 22 to 201 with a median of 74.^18, 70, 74, 84, 88^ All studies used the PSYRATS AH scoring system and were carried out across three different countries and continents: UK, Canada, and China. AVATAR therapy showed a medium and statistically significant effect size of −0.425 (95% CI, [−0.601, - 0.250], Z = −4.741, p < 0.001) (Fig. 4). Effect sizes were highly consistent across studies, showing no significant heterogeneity (Q= 2.649, I^2^ = 0%).

**Fig. 4.**
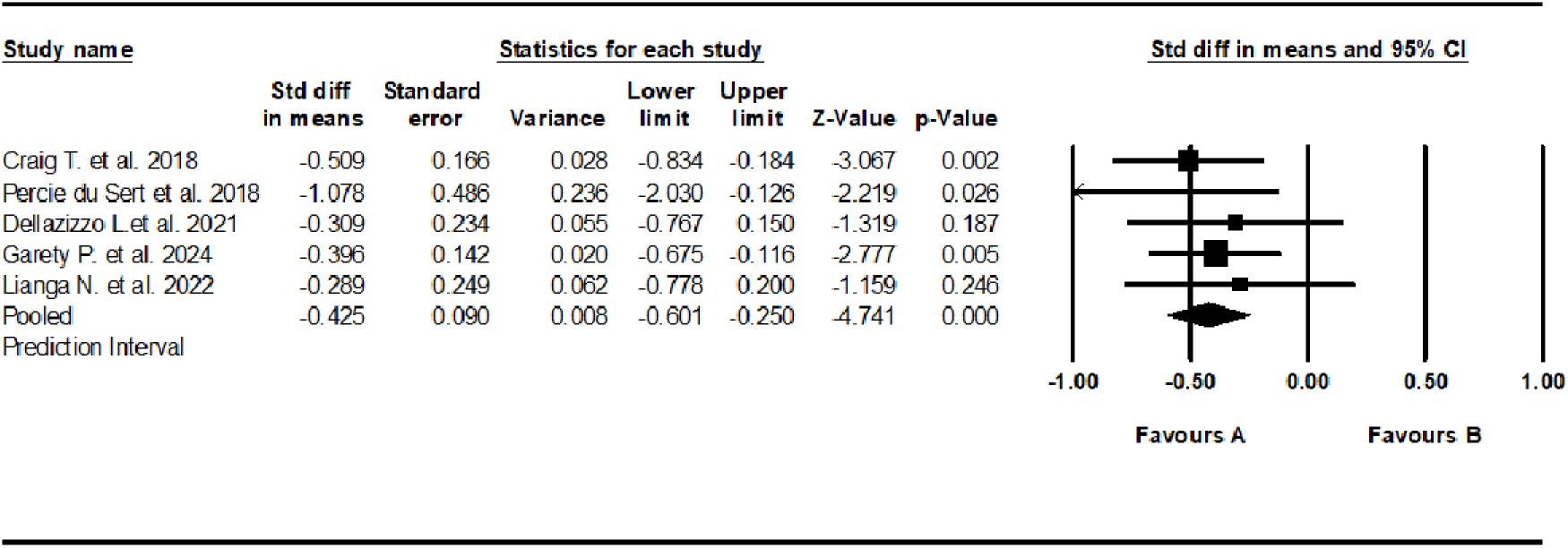
Forest plot of standardized mean differences (SMD) between post-intervention AVH scores in those who received AVATAR therapy for AVHs (A), versus the control group (B). Negative effect sizes indicate lower AVH scores in the intervention group compared to the control group.

### Cognitive behavioural therapy

Eight studies tested the use of CBT and had sample sizes ranging from 26 to 116, with a median of 35.^51, 53, 54, 61, 62, 64^, ^71^AVH scoring systems varied between studies, including both HPSVQ and PSYRATS AH. Studies were carried out across six different countries including: Pakistan, UK, Canada, USA, Norway and Turkey. CBT showed a medium and statistically significant effect size of −0.396 (95% CI, [−0.764, −0.027], Z = −2.103, p = 0.035) (Fig. 5). Effect sizes were highly inconsistent across studies, showing moderate and statistically significant heterogeneity (Q= 19.624, I^2^ = 64%, p = 0.006).

**Fig. 5.**
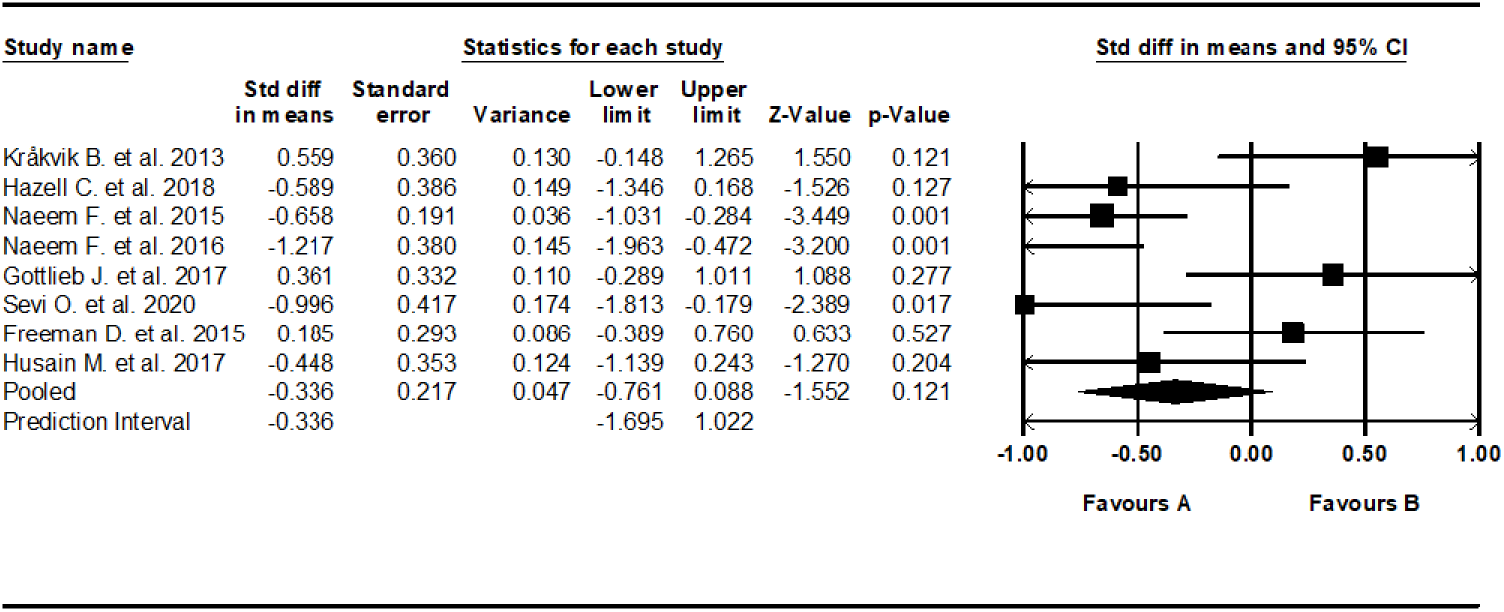
Forest plot of standardized mean differences (SMD) between post-intervention AVH scores in those who received CBT for AVHs, versus the control group. Negative effect sizes indicate lower AVH scores in the intervention group compared to the control group.

### Repetitive transcranial magnetic stimulation

Nine studies tested the use of rTMS and had sample sizes ranging from 27 to 64, with a median of 36.^17, 25, 50, 52, 55, 57, 58, 65, 86^ All studies used the AHRS and were carried out across six different countries: Czech Republic, The Netherlands, France, India, Japan, and China. rTMS showed a small and not statistically significant effect size of −0.267 (95% CI, [−0.788 to 0.255], Z = −1.003, p = 0.316) (Fig. 6). Effect sizes were highly inconsistent across studies, showing substantial and statistically significant heterogeneity (Q = 50.842, I^2^ = 84%, p< 0.001).

**Fig. 6.**
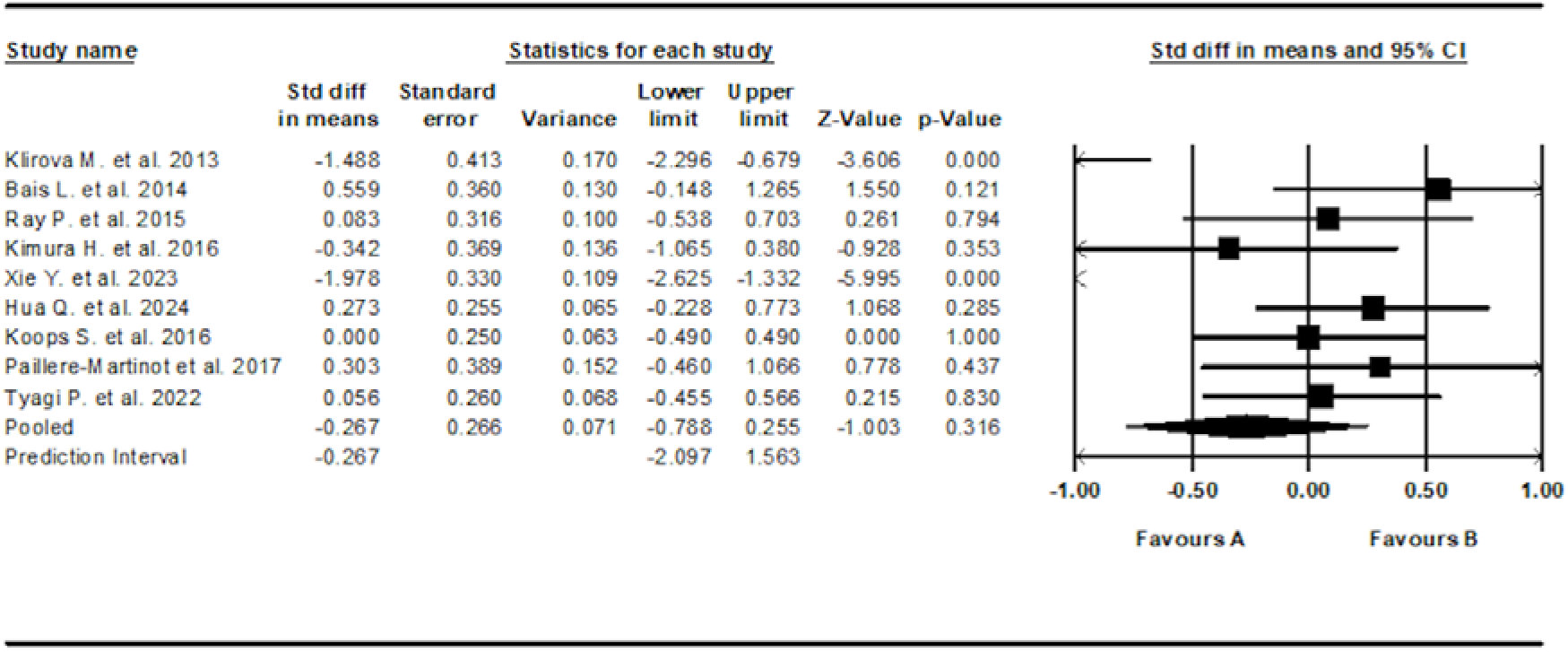
Forest plot of standardized mean differences (SMD) between post-intervention AVH scores in those who received rTMS for AVHs (A), versus the control group (B). Negative effect sizes indicate lower AVH scores in the intervention group compared to the control group.

### tDCS

Eleven studies tested the use of tDCS and had sample sizes ranging from 7 to 50, with a median of 13.^49, 56, 60, 68, 69, 72, 73, 76, 77, 80, 83^ Scoring systems used varied between studies, including both the AHRS and PANSS AH. Studies were carried out across eight different countries including: USA, France, India, Taiwan, The Netherlands, Brazil, Ireland and Canada. tDCS showed a very small and not statistically significant effect size of −0.141(95% CI, [−0.367, 0.086], Z = −1.085, p = 0.278) (Fig. 7). Effect sizes were inconsistent across studies, showing moderate, but not statistically significant heterogeneity (Q= 16.105, I^2^ = 34%, p= 0.097).

**Fig. 7.**
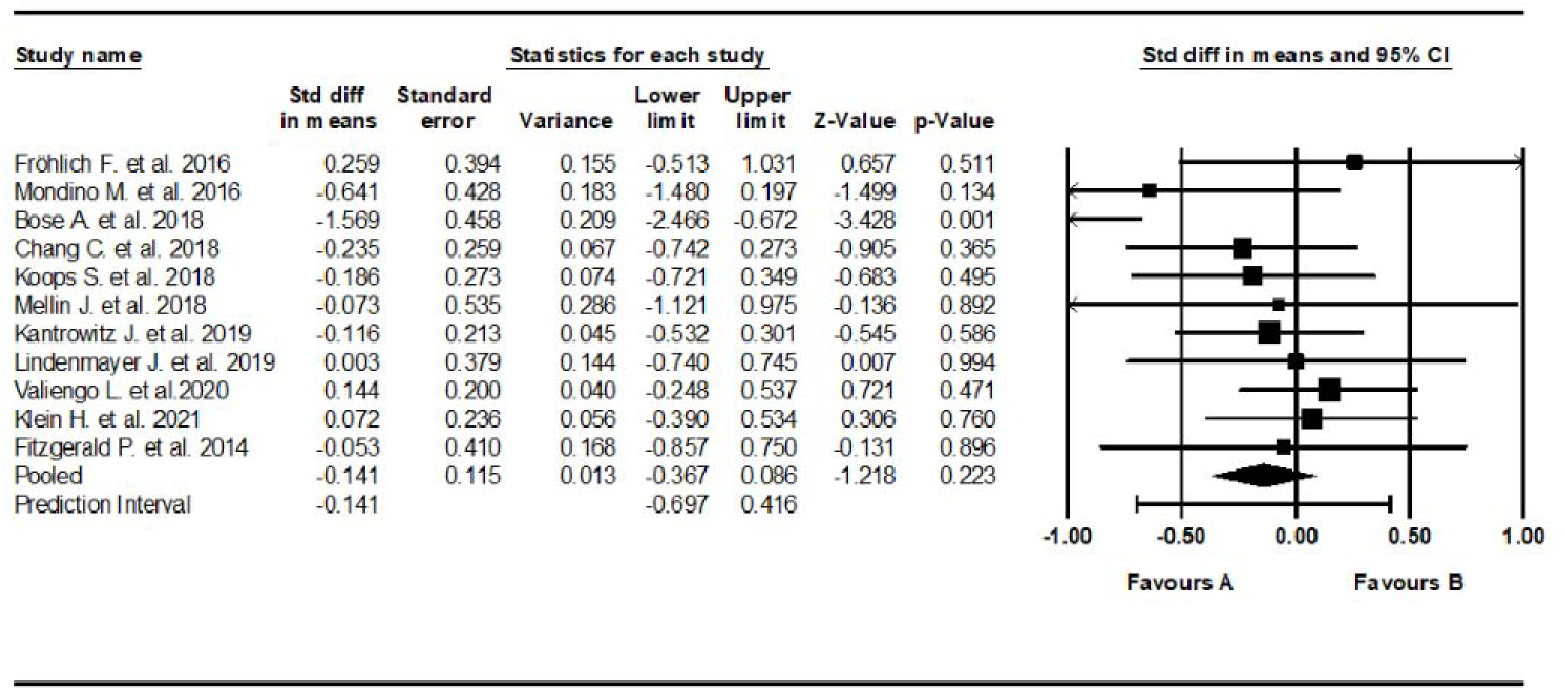
Forest plot of standardized mean differences (SMD) between post-intervention AVH scores in those who received tDCS for AVHs (A), versus the control group (B). Negative effect sizes indicate lower AVH scores in the intervention group compared to the control group.

### Transcranial alternating current stimulation

The study by Zhang M. et al. reported a medium size, non-significant effect, favouring those in the control group (Hedge’s g = 0.569 (95% CI, [−0.510, 1.649], p = 0301, Z = 1034).

### Smartphone app

Two studies trialled a smartphone app intervention. The study by Bell I. et al. trialled blended coping-focused therapy using ecological momentary assessment and intervention via a smartphone app. They found a medium, but not statistically significant effect size favouring the intervention (Hedge’s g = −0.558, 95% CI [−1.22, 0.127], p = 0.11, z = −1.597). The study by Jongeneel A. et al.^89^ trialled the use of the smartphone app, Temstem, which consists of two language games: ‘Silencing’ to improve control over voices, and ‘Challenging’ to reduce vividness and emotionality of voices. No significant difference was observed between those using the two language games on the Temstem app, and those in the control group (Hedge’s g = −0.092, 95% CI [−0.507, 0.324], p = 0.666, z = −0.432).

### Psychological online intervention

Lüdtke et al.^79^ found that partaking in a POI resulted in no significant difference in AVHs compared to the control group (Hedge’s g = −0.089, 95% CI [−0.672, 0.493], p = 0.763, z = −0.301).

### Acceptance and commitment therapy

Two studies trialled ACT. Shawyer F. et al. found that ACT demonstrated no significant difference in AVHs compared to the control group (Hedge’s g = 0.013, 95% CI [−3.595, −2.189], p = 0.00, z = −8.07). In contrast, El Ashry A. et al. found that ACT demonstrated a very large and statistically significant effect size, favouring the intervention (Hedge’s g = −2.891, 95% CI [−3.595, −2.189], p = 0.00, z = −8.07).

### Music therapy

Rast tonality MT, explored by Pinar S. et al., found a large, statistically significant effect size favouring the control group, who did not listen to any music whilst in hospital (Hedge’s g = 0.972, 95% CI [0.189, 1.756], p = 0.015, z = 2.433).

### Relating therapy

Hayward M. et al.^63^ found that RT showed a medium, but not statistically significant effect size, favouring the intervention (Hedge’s g = −0.466, 95% CI [−1.245, 0.313], p = 0.241, z = −1.172).

### Counselling

Schnackenberg J. et al.^66^ found that experienced focused counselling, had a very small and non-significant effect size, favouring the intervention (Hedge’s g = −0.092, 95% CI [−0.507, 0.896], p = 0.662, z = 0.437).

### Rehabilitation (attention training programme)

The study by López-Luengo B. et al. found that rehabilitation in the form of an attention training programme produced a large, but not statistically significant effect size, favouring the intervention (Hedge’s g = −0.818, 95% CI [−3.593, −2.189], p = 0.116, z = −1.571).

### Referral to community psychologist

Solar A. et al. found that referring participants to a community psychologist resulted in a medium and non-significant effect size, favouring the control, which involved TAU (standard hospital care, with no direct psychologist appointment, but could self-request a referral) (Hedge’s g = 0.376, 95% CI [−0.598, 1.350], p = 0.449, z = 0.757).

### Group mindfulness-based intervention

Chadwick P. et al. found that receiving a group mindfulness-based intervention resulted in no significant difference in AVHs compared to the control group (Hedge’s g = −0.065, 95% CI [−0.442, 0.312], p = 0.736, z = −0.337).

## Discussion

### Summary of findings

This systematic review and meta-analysis summarised evidence from the last decade on fourteen different non-pharmacological interventions for schizophrenia and related disorders, through an evaluation of RCT studies. AVATAR therapy demonstrated the most favourable results for treating AVHs and was highly consistent across all studies. The larger sample sizes and geographical spread of RCTs exploring AVATAR therapy increase the generalisability of results. Overall, AVATAR therapy stands out as an excellent candidate for future clinical implementation for treating AVHs. CBT had promising, and statistically significant results of moderate effect size for improving AVHs, however substantial heterogeneity was observed between studies. This may be due to the differences in methodologies employed in different trials, such as differences in CBT techniques, treatment duration and therapist expertise. Non-invasive brain stimulation techniques including rTMS, tDCS, and tACS all failed to produce statistically significant outcomes for the treatment of AVHs. However, rTMS had large variability in results across different studies, suggesting that differences in methodologies, potentially concerning anatomical location, and exact rTMS protocol, along with possible differences in patient populations, may be impacting the results. In contrast tDCS studies showed no significant heterogeneity, highlighting that more studies consistently did not show an improvement in AVHs post tDCS administration. This conclusion is reinforced by the fact that a large number of studies (11), conducted across the globe, were included in the subgroup analysis, increasing the diversity of samples, and therefore generalisability of the results. ACT demonstrated variable effectiveness at treating AVHs across the two studies. However, despite limited evidence for ACT, the study by El Ashry A. et al. had a moderate sample size, a very high QualSyst bias assessment score suggestive of low risk of bias, and was the only study carried out in Africa, a continent frequently underrepresented in research. As therapy is culturally influenced,^90^ demonstrating this effect in a non-Western setting could have great implication for effectively treating AVHs in these contexts. However, the contrasting results found by Shawyer F. et al.^67^ demonstrate a need for more trials refining the most effective method for delivering ACT. Rehabilitation, counselling, RT, and referral to a community psychologist all showed improvement in AVHs post-treatment, but this was not statistically significant.

Receiving a group mindfulness-based intervention did not show statistical improvement in reducing AVHs. Smartphone apps and POI also failed to show statistical improvement in reducing AVHs, potentially due to the way in which these treatments were delivered, suggesting that online platforms may benefit from future refinements before they can be considered for clinical applications. Lastly, MT demonstrated significantly better AVH outcomes in the control group post-treatment, who did not listen to any music during their hospital stay. However, at follow up, there was an improvement in AVHs in those who received MT, highlighting the value of long-term and follow-up RCTs for the treatment of AVHs.

### Future research directions and clinical implications

By comparing these findings to existing systematic reviews and meta-analyses, we can assess the consistency, divergence, and potential advancements in understanding the efficacy of treatments for AVHs in schizophrenia spectrum disorders. A previous systematic review by Dellazizzo L. et al. explored relational-based therapies for AVHs, including both AVATAR therapy and CBT. Whilst CBT focuses on changing the person’s cognition surrounding their AVHs by, for instance, altering their belief that they must obey their AVHs, AVATAR therapy works on improving the relationship between the person and their AVHs, aiming to give the patient a sense of control. The present meta-analysis found that AVATAR therapy had a moderate size and statistically significant effect favouring the intervention.

Dellazizzo L. et al.’s reported large or very large effect sizes for AVATAR therapy. The difference in effect size may be partially explained by the fact that the previous review included studies ranging from very low to high risk of bias, whereas only studies with low risk of bias were included in the present review. The reliability of our findings is reinforced by the low heterogeneity for AVATAR therapy (I ^2^= 0.00%), making the conclusion valuable for evidence-based clinical practice. Future research should continue to focus on synthesising high-quality evidence for AVATAR therapy. RCTs should continue evaluating its effectiveness, and work towards implementing AVATAR therapy into clinical practice.

According to the National Institute for Health and Care Excellence (NICE), CBT is currently a recognised treatment for schizophrenia and psychosis and is included in the NICE guidelines as a best-practice treatment recommendation. However, evidence from Dellazizzo L. et al.’s systematic review reports that across the five studies included, all but one case series failed to show any significant effects of CBT in reducing PSYRATS AH scores. Similarly, the high heterogeneity across the eight studies included in this meta-analysis (I^2^ = 64%) warrants cautious interpretation of the moderate and statistically significant effect size found in favour of CBT. This prompts an exploration of the different methodologies and samples used in trials for CBT. More high-quality research following the methodology of successful trials is needed to increase the reliability of results and to refine the most effective way to deliver CBT. Ultimately, these results suggest that CBT may lack the evidence to justify its utilisation in clinical practice for AVHs, and guidelines should be revised to specify which patient groups may benefit from CBT, as it is currently a general recommendation for those with schizophrenia or psychosis.^91^ However, if AVHs are a main, distressing, and debilitating symptom for an individual, CBT may not be the appropriate treatment for them, and guidelines should reflect symptomology rather than general diagnoses, particularly in the context of schizophrenia spectrum disorders. Additionally, alternative therapies should be considered for best-practice guidelines, such as AVATAR therapy which has shown greater evidence for effectiveness against AVHs compared to CBT.

ACT is another variant of CBT, which focuses on helping individuals to reduce the distress caused by voices. A systematic review by Yıldız E. included three studies which explored AVH outcomes post ACT. The study found that two out of the three studies reported significant improvements in AVHs after receiving ACT. This variability in effectiveness is also present in our results. Given the limited number of RCTs for ACT, this intervention remains under explored and future trials must be conducted to draw reliable conclusions.

With regards to non-invasive brain stimulation techniques, a systematic review and meta-analysis of ten studies by Otani V. et al., exploring the use of rTMS for treatment of auditory hallucinations in refractory schizophrenia, found a moderate, statistically significant effect size favouring rTMS (Hedge’s g = 0.49, 95% CI [0.11, 0.88], p = 0.011).

Conversely, we found a small, not statistically significant effect size (Hedge’s g = −0.267, [95% CI, −0.788, 0.255], p = 0.316), in favour of rTMS. Multiple factors may have contributed to this difference. Firstly, it is important to note that there was no crossover in the papers included in this meta-analysis and Otani V. et al.’s meta-analysis, which focused on papers from the years 2000 to 2011. Additionally, Otani V. et al. included studies which used low frequency rTMS and rTMS applied only on the left temporoparietal cortex (leTPC).^19^ Studies in the current meta-analysis included rTMS applied at any anatomical location, including the leTPC, left temporo-parietal junction (TPJ), right TPJ, and personalised localisation of the language perception area. Additionally, our study included high-frequency stimulation, continuous theta-burst stimulation, and low frequency stimulation. These differences may indicate that low-frequency rTMS applied exclusively to the leTPC may be more effective in treating AVHs, highlighting a need for more trials exploring this, and an up-to date meta-analysis on studies with this inclusion criterion. However, the meta-analysis by Otani V. et al. did not evaluate each paper for risk of bias prior to inclusion, which may have also affected the results.

Furthermore, a previous systematic review and meta-analysis of sixteen studies exploring tDCS by Cheng P. et al., found a moderate and statistically significant effect size in favour of tDCS (SMD = 0.36, 95 % CI [0.02, 0.70]). Although tDCS is also a non-invasive brain stimulation technique, it utilises an electrical current applied across the patients’ scalp,^94^ versus the use of pulsed magnetic fields in rTMS.^95^ tDCS also has greater potential for use in clinical practice due to being more cost effective and easily accessible than rTMS.^96^ However, Cheng P. et al. observed substantial heterogeneity across the included studies (I^2^ = 65%), and statistical significance did not survive subsequent sensitivity analyses. Similarly, our meta-analysis found a small and not statistically significant effect size in favour of tDCS (−0.141, 95% CI, [−0.367 to 0.086]), with no significant heterogeneity, reinforcing the reliability of the negative result. Another systematic review of nine studies exploring tDCS by Rashidi S. et al. found that tDCS was no better than sham tDCS in reducing AVH in 3 studies, and that tDCS had a significant effect in reducing AVHs in the remaining six studies.^21^ The variability in results across systematic reviews suggests that more high quality RCTs exploring tDCS may be needed to confirm effectiveness.

### Strengths and limitations

This systematic review and meta-analysis has a number of strengths. Firstly, the inclusion of a large sample of 45 studies with a total of 2,314 participants increases the statistical power of the meta-analysis and provides a comprehensive summary of evidence from the last decade. Secondly, PRISMA guidelines were followed, and a robust bias assessment was carried out on each included study, along with assessment of a funnel plot for publication bias. The funnel plot did not demonstrate obvious publication bias, suggesting that a range of studies with both positive and negative findings were published.^97^ This increases the reliability of our results. Since only high-quality evidence was included, results can be used to influence future research priorities, with potential to influence clinical practice and healthcare policy. Furthermore, some subgroup analyses had low heterogeneity, particularly for AVATAR therapy. This enables a definite interpretation of effect size,^40^ which can provide a clear guide for future research plans and clinical applications.

This systematic review and meta-analysis also has some limitations, which must be addressed. Firstly, eleven eligible studies had to be excluded for missing data regarding pre and post treatment AVH scores, despite contacting authors with data requests. Missing data can introduce bias, as negative or non-significant results may have been omitted, leading to a potential overestimation of effect sizes.^98^ Secondly, some studies included follow up data, which were not explored in this study. This prevents consideration for long-term impacts of treatments, as some effects may either diminish, remain stable, or strengthen over time, which is important to consider for clinical implementation of treatments.^99^ This highlights a need for future RCTs to collect follow up data, and meta-analyses to stratify and analyse follow up data. Thirdly, some subgroups showed substantial and significant heterogeneity in their results, particularly for rTMS and CBT. This makes findings harder to interpret, as high heterogeneity may weaken the generalisability and reliability of pooled effect sizes. For example, findings may reflect study specific factors, rather than true treatment efficacy.^100^ Furthermore, all studies trialled the interventions on top of existing treatment regimens, therefore isolated effects of the intervention on AVHs are impossible to determine, reducing internal validity.^101^ However, this may strengthen external validity by providing a better reflection for the utilisation of these interventions in real-world clinical and community settings.^101^ The presence of some participants who did not have SSDs, but instead had mood or personality disorders with AVHs, may limit how reliably conclusions can be drawn about the treatment of AVHs in SSDs specifically. To address this, future meta-analyses should have rigid inclusion criteria for diagnoses. However, this meta-analysis included such studies to ensure a comprehensive overview of all recent RCTs for treatments of AVHs, where the majority of participants had SSDs. Additionally, studies support that AVHs may be phenomenologically similar across these conditions^102, 103^ and respond similarly to established treatments such as antipsychotics.^102^ Lastly, not all studies had a placebo, inactive, or TAU control group. For instance, the study exploring AVATAR therapy by Liang N. et al.^84^ utilised an active control group which received CBT, on top of TAU. This makes the true effect of the intervention unclear as the two interventions were compared head-to-head.

### Conclusion

This systematic review and meta-analysis highlights that AVATAR therapy has the strongest base of evidence for treating AVHs in SSDs. It is of clinical priority to continue large scale RCTs for AVATAR therapy and to integrate it into treatment guidelines in the near future.

Conversely, CBT requires standardisation to improve reliability with regards to its effectiveness in treating AVHs, and this finding underscores a need for widely implemented clinical guidelines, such as the NICE guidelines, to shift from diagnosis-based to symptom-driven recommendations for SSDs. Notably, ACT remains a promising approach for reducing distress associated with AVHs, requiring further research and refinement for clinical use. There is also a need for more geographically diverse RCTs, and to build on studies such as El. Ashry et al.’s which tested ACT in a non-Western context. Lastly, non-invasive brain stimulation techniques require further trials before considering them for clinical adoption.

## Author contributions

MC and NS conceived the study, carried out the data collection, bias analysis, statistical analyses and interpretation. MC wrote the first draft, and MC and NS revised and approved the final draft.

## Data availability

Data availability is not applicable to this article as no new data were created or analysed in this study.

## Affiliations

Both authors are affiliated with the Department of Clinical Neurosciences, Brighton and Sussex Medical School. Trafford Centre, University of Sussex, Falmer. BN1 9RY. UK. All work was carried out in the UK.

**Corresponding author:** Natasha Sigala

## Supplementary materials

1. PRISMA-P (or equivalent) table
2. Search strings used for various platforms such as MEDLINE, Scopus etc.
3. Number of each intervention type included
4. List of all included papers
5. Publication bias: funnel plot
6. Quality Assessment

---

**Table S1.**
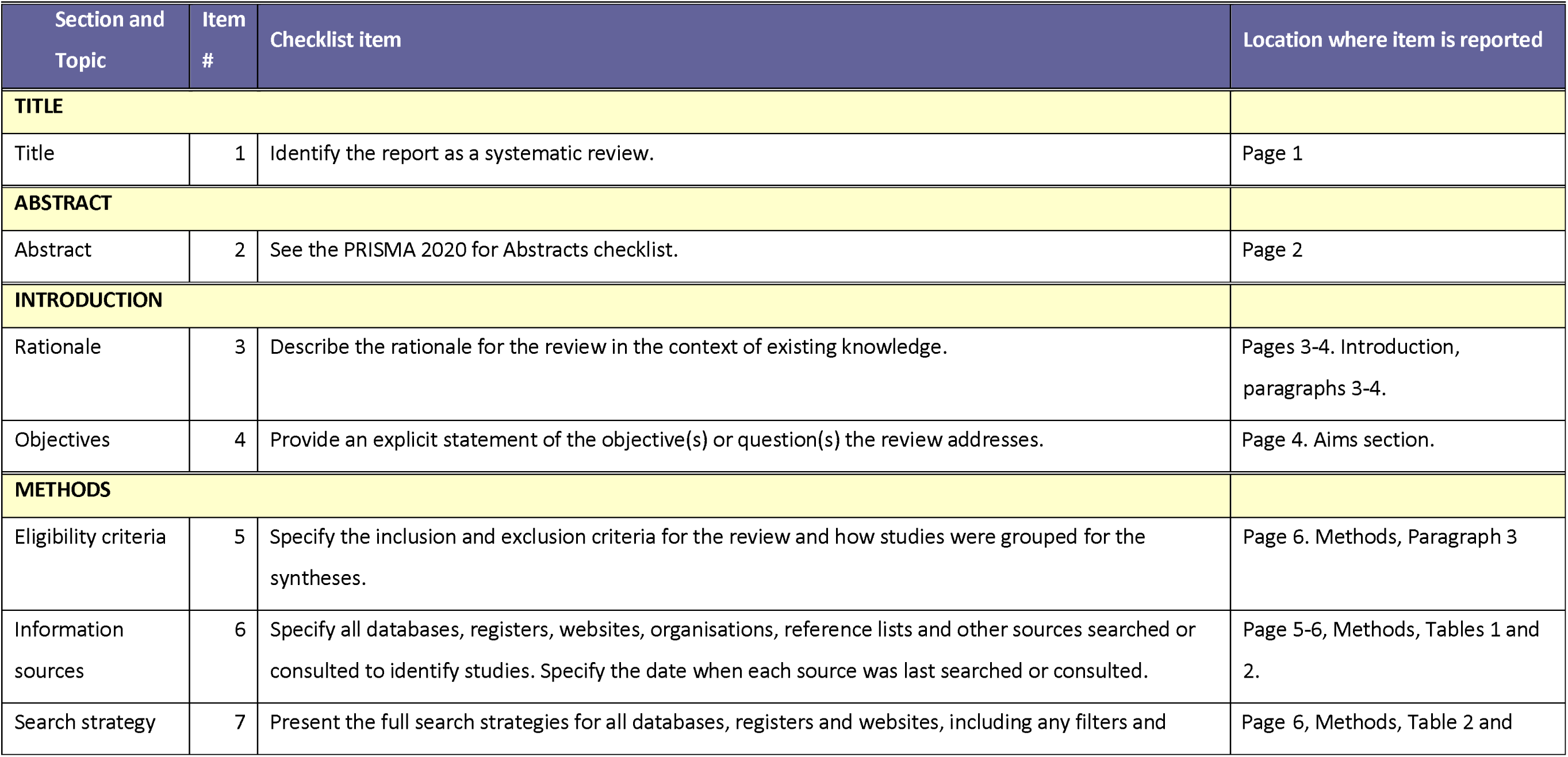
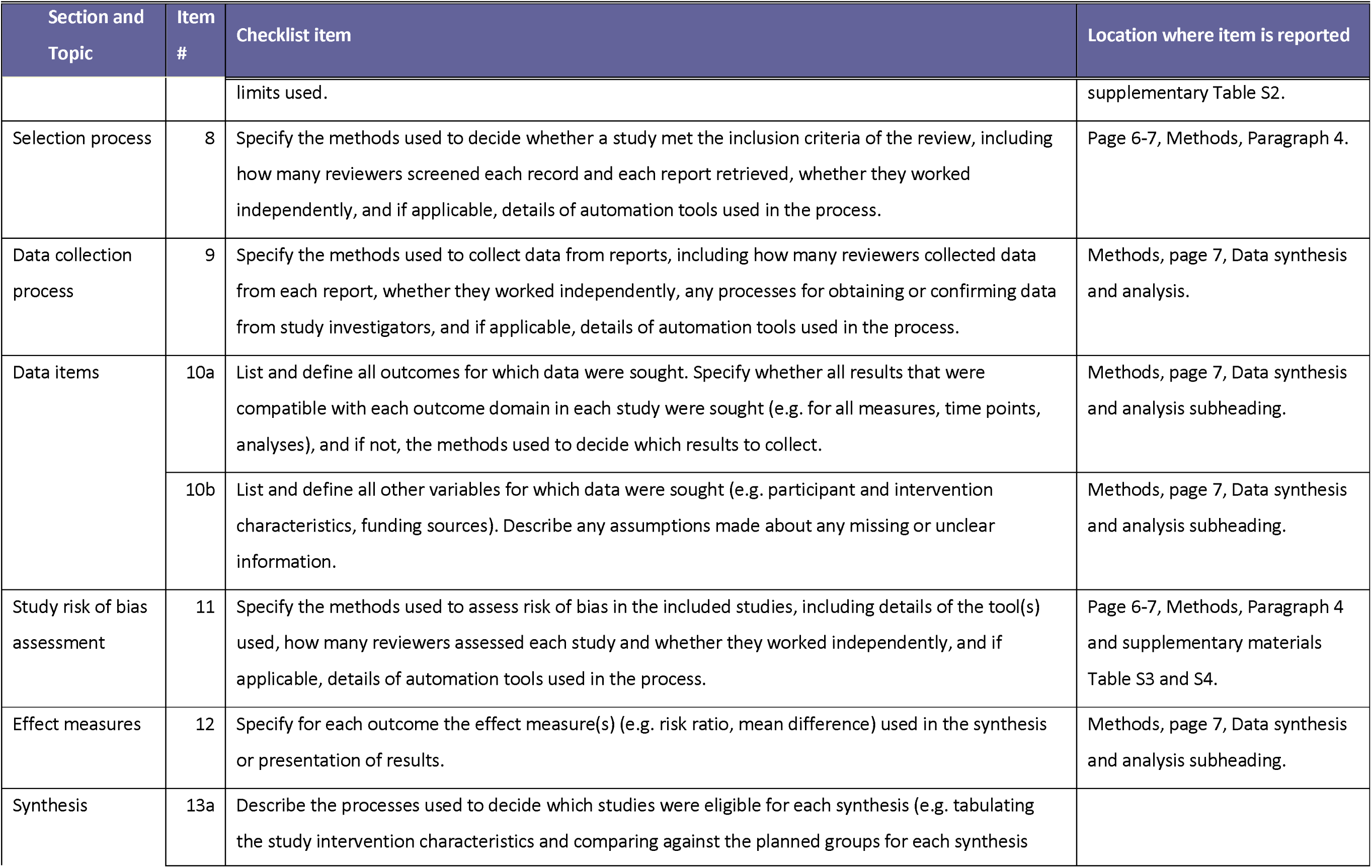
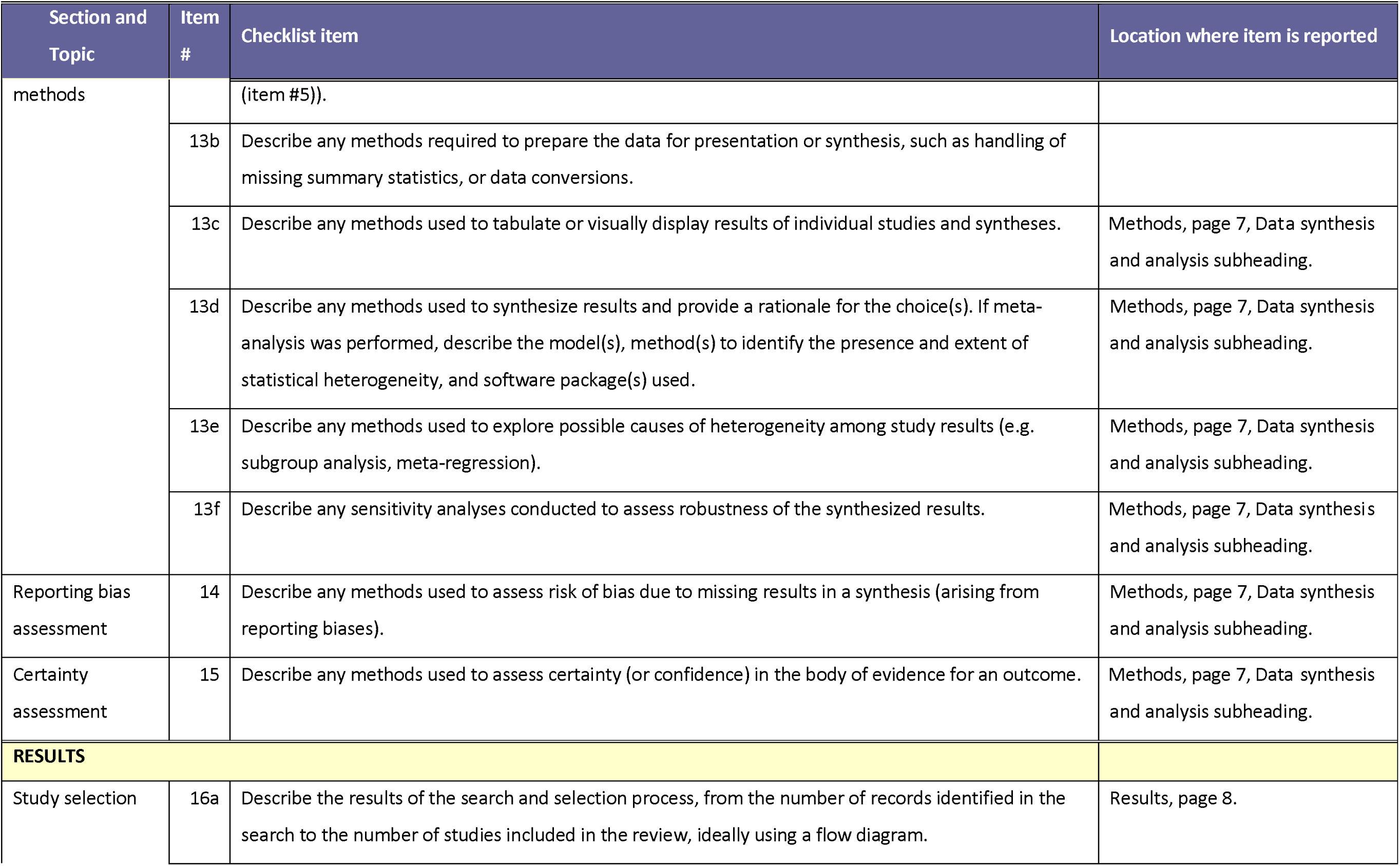
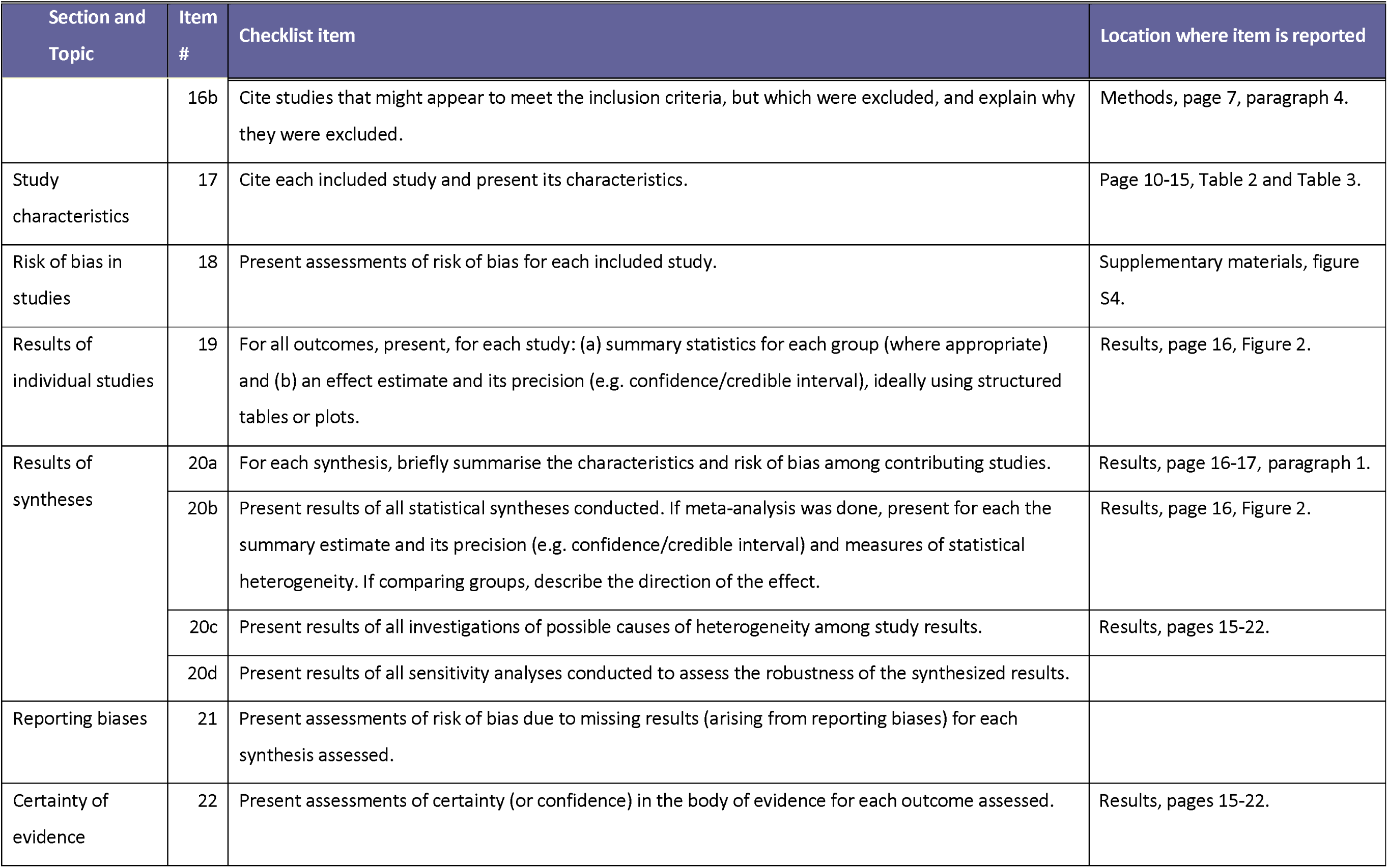
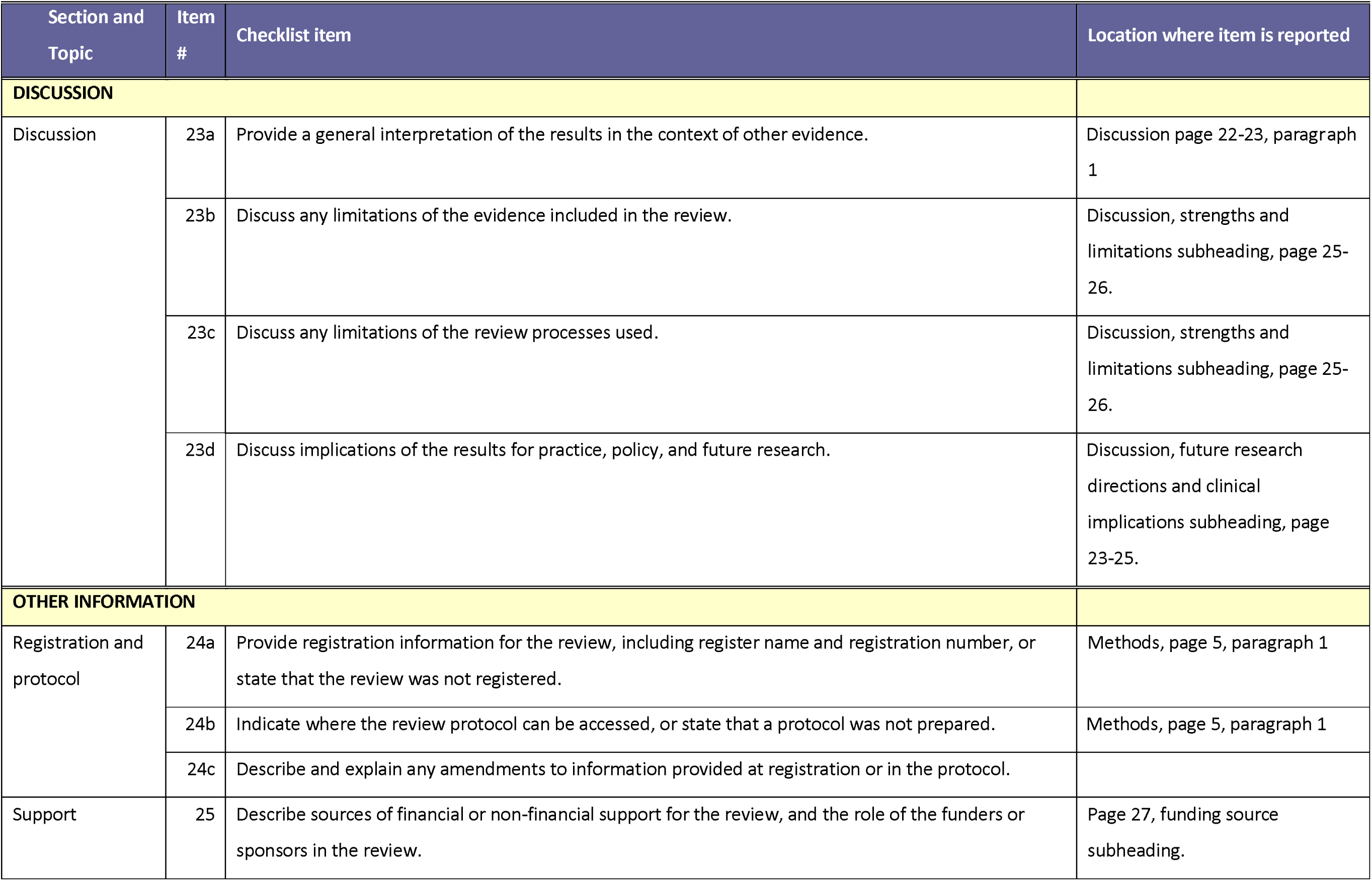
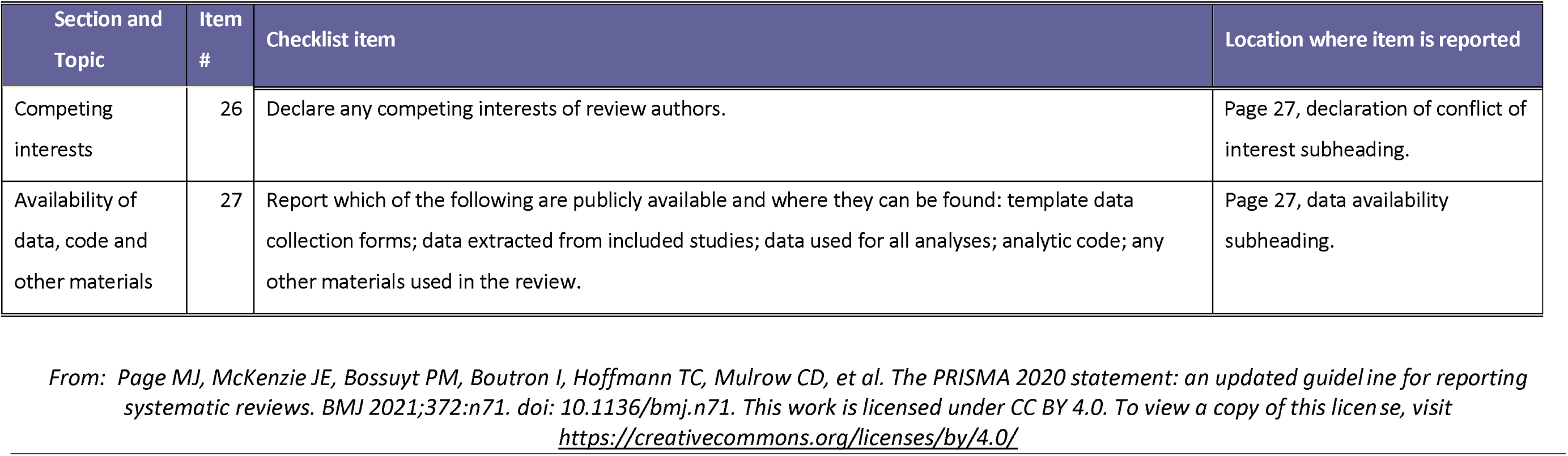
PRISMA 2020 checklist: Key items to report in a systematic review.

| Section and Topic | Item # | Checklist item | Location where item is reported |
| --- | --- | --- | --- |
| **TITLE** |  |  |  |
| Title | 1 | Identify the report as a systematic review. | Page 1 |
| **ABSTRACT** |  |  |  |
| Abstract | 2 | See the PRISMA 2020 for Abstracts checklist. | Page 2 |
| **INTRODUCTION** |  |  |  |
| Rationale | 3 | Describe the rationale for the review in the context of existing knowledge. | Pages 3-4. Introduction, paragraphs 3-4. |
| Objectives | 4 | Provide an explicit statement of the objective(s) or question(s) the review addresses. | Page 4. Aims section. |
| **METHODS** |  |  |  |
| Eligibility criteria | 5 | Specify the inclusion and exclusion criteria for the review and how studies were grouped for the syntheses. | Page 6. Methods, Paragraph 3 |
| Information sources | 6 | Specify all databases, registers, websites, organisations, reference lists and other sources searched or consulted to identify studies. Specify the date when each source was last searched or consulted. | Page 5-6, Methods, Tables 1 and 2. |
| Search strategy | 7 | Present the full search strategies for all databases, registers and websites, including any filters and | Page 6, Methods, Table 2 and |
|  |  | limits used. | supplementary Table S2. |
| Selection process | 8 | Specify the methods used to decide whether a study met the inclusion criteria of the review, including how many reviewers screened each record and each report retrieved, whether they worked independently, and if applicable, details of automation tools used in the process. | Page 6-7, Methods, Paragraph 4. |
| Data collection process | 9 | Specify the methods used to collect data from reports, including how many reviewers collected data from each report, whether they worked independently, any processes for obtaining or confirming data from study investigators, and if applicable, details of automation tools used in the process. | Methods, page 7, Data synthesis and analysis. |
| Data items | 10a | List and define all outcomes for which data were sought. Specify whether all results that were compatible with each outcome domain in each study were sought (e.g. for all measures, time points, analyses), and if not, the methods used to decide which results to collect. | Methods, page 7, Data synthesis and analysis subheading. |
|  | 10b | List and define all other variables for which data were sought (e.g. participant and intervention characteristics, funding sources). Describe any assumptions made about any missing or unclear information. | Methods, page 7, Data synthesis and analysis subheading. |
| Study risk of bias assessment | 11 | Specify the methods used to assess risk of bias in the included studies, including details of the tool(s) used, how many reviewers assessed each study and whether they worked independently, and if applicable, details of automation tools used in the process. | Page 6-7, Methods, Paragraph 4 and supplementary materials Table S3 and S4. |
| Effect measures | 12 | Specify for each outcome the effect measure(s) (e.g. risk ratio, mean difference) used in the synthesis or presentation of results. | Methods, page 7, Data synthesis and analysis subheading. |
| Synthesis | 13a | Describe the processes used to decide which studies were eligible for each synthesis (e.g. tabulating the study intervention characteristics and comparing against the planned groups for each synthesis |  |
| methods |  | (item #5)). |  |
|  | 13b | Describe any methods required to prepare the data for presentation or synthesis, such as handling of missing summary statistics, or data conversions. |  |
|  | 13c | Describe any methods used to tabulate or visually display results of individual studies and syntheses. | Methods, page 7, Data synthesis and analysis subheading. |
|  | 13d | Describe any methods used to synthesize results and provide a rationale for the choice(s). If meta-analysis was performed, describe the model(s), method(s) to identify the presence and extent of statistical heterogeneity, and software package(s) used. | Methods, page 7, Data synthesis and analysis subheading. |
|  | 13e | Describe any methods used to explore possible causes of heterogeneity among study results (e.g. subgroup analysis, meta-regression). | Methods, page 7, Data synthesis and analysis subheading. |
|  | 13f | Describe any sensitivity analyses conducted to assess robustness of the synthesized results. | Methods, page 7, Data synthesis and analysis subheading. |
| Reporting bias assessment | 14 | Describe any methods used to assess risk of bias due to missing results in a synthesis (arising from reporting biases). | Methods, page 7, Data synthesis and analysis subheading. |
| Certainty assessment | 15 | Describe any methods used to assess certainty (or confidence) in the body of evidence for an outcome. | Methods, page 7, Data synthesis and analysis subheading. |
| **RESULTS** |  |  |  |
| Study selection | 16a | Describe the results of the search and selection process, from the number of records identified in the search to the number of studies included in the review, ideally using a flow diagram. | Results, page 8. |
|  | 16b | Cite studies that might appear to meet the inclusion criteria, but which were excluded, and explain why they were excluded. | Methods, page 7, paragraph 4. |
| Study characteristics | 17 | Cite each included study and present its characteristics. | Page 10-15, Table 2 and Table 3. |
| Risk of bias in studies | 18 | Present assessments of risk of bias for each included study. | Supplementary materials, figure S4. |
| Results of individual studies | 19 | For all outcomes, present, for each study: (a) summary statistics for each group (where appropriate) and (b) an effect estimate and its precision (e.g. confidence/credible interval), ideally using structured tables or plots. | Results, page 16, Figure 2. |
| Results of syntheses | 20a | For each synthesis, briefly summarise the characteristics and risk of bias among contributing studies. | Results, page 16-17, paragraph 1. |
|  | 20b | Present results of all statistical syntheses conducted. If meta-analysis was done, present for each the summary estimate and its precision (e.g. confidence/credible interval) and measures of statistical heterogeneity. If comparing groups, describe the direction of the effect. | Results, page 16, Figure 2. |
|  | 20c | Present results of all investigations of possible causes of heterogeneity among study results. | Results, pages 15-22. |
|  | 20d | Present results of all sensitivity analyses conducted to assess the robustness of the synthesized results. |  |
| Reporting biases | 21 | Present assessments of risk of bias due to missing results (arising from reporting biases) for each synthesis assessed. |  |
| Certainty of evidence | 22 | Present assessments of certainty (or confidence) in the body of evidence for each outcome assessed. | Results, pages 15-22. |
| **DISCUSSION** |  |  |  |
| Discussion | 23a | Provide a general interpretation of the results in the context of other evidence. | Discussion page 22-23, paragraph 1 |
|  | 23b | Discuss any limitations of the evidence included in the review. | Discussion, strengths and limitations subheading, page 25-26. |
|  | 23c | Discuss any limitations of the review processes used. | Discussion, strengths and limitations subheading, page 25-26. |
|  | 23d | Discuss implications of the results for practice, policy, and future research. | Discussion, future research directions and clinical implications subheading, page 23-25. |
| **OTHER INFORMATION** |  |  |  |
| Registration and protocol | 24a | Provide registration information for the review, including register name and registration number, or state that the review was not registered. | Methods, page 5, paragraph 1 |
|  | 24b | Indicate where the review protocol can be accessed, or state that a protocol was not prepared. | Methods, page 5, paragraph 1 |
|  | 24c | Describe and explain any amendments to information provided at registration or in the protocol. |  |
| Support | 25 | Describe sources of financial or non-financial support for the review, and the role of the funders or sponsors in the review. | Page 27, funding source subheading. |
| Competing interests | 26 | Declare any competing interests of review authors. | Page 27, declaration of conflict of interest subheading. |
| Availability of data, code and other materials | 27 | Report which of the following are publicly available and where they can be found: template data collection forms; data extracted from included studies; data used for all analyses; analytic code; any other materials used in the review. | Page 27, data availability subheading. |

*From: Page MJ, McKenzie JE, Bossuyt PM, Boutron I, Hoffmann TC, Mulrow CD, et al. The PRISMA 2020 statement: an updated guideline for reporting systematic reviews. BMJ 2021;372:n71. doi: 10.1136/bmj.n71. This work is licensed under CC BY 4.0. To view a copy of this license, visit https://creativecommons.org/licenses/by/4.0/*

**Table S2.** Full search strings used for each database.

| Database | Search String |
| --- | --- |
| PubMed | ("auditory hallucination*" OR "hearing voices" OR "voice-hearing" OR "auditory hallucinations" OR<br>("voices" AND "psychosis"))<br>AND<br>("Schizophrenia"[MeSH Terms] OR schizophren* OR "psychotic disorders" OR "psychosis" OR |
|  | "schizoaffective disorder")<br>AND<br>("Treatment Outcome"[MeSH Terms] OR "therapy" OR "treatment efficacy" OR "antipsychotic agents"[MeSH Terms] OR "Clozapine" OR "Haloperidol" OR "Olanzapine" OR "Risperidone" OR "Aripiprazole" OR "CBT" OR "Cognitive Behavioral Therapy" OR "cognitive remediation therapy" OR "psychoeducation" OR "family therapy" OR "supportive therapy" OR "electroconvulsive therapy" OR "rTMS" OR "transcranial magnetic stimulation" OR "tDCS" OR "transcranial direct current stimulation" OR "Deep Brain Stimulation" OR "DBS" OR "virtual reality therapy" OR "VR therapy" OR "avatar therapy" OR "digital intervention" OR "mHealth" OR "eHealth" OR "psychedelic-assisted psychotherapy" OR "psilocybin") |
| **Embase** | ("auditory hallucination*" OR "hearing voices" OR "voice-hearing" OR "hallucinations, auditory" OR ("voices" AND "psychosis"))<br>AND<br>("schizophrenia" OR schizophren* OR "psychotic disorders" OR "psychosis" OR "schizoaffective disorder")<br>AND<br>("treatment outcome" OR "treatment efficacy" OR "therapy" OR "antipsychotic agents" OR "Clozapine" OR "Haloperidol" OR "Olanzapine" OR "Risperidone" OR "Aripiprazole" OR "Cognitive Behavioral Therapy" OR "CBT" OR "cognitive remediation therapy" OR "psychoeducation" OR "family therapy" OR "supportive therapy" OR "electroconvulsive therapy" OR "rTMS" OR "transcranial magnetic stimulation" OR "tDCS" OR "transcranial direct current stimulation" OR "Deep Brain Stimulation" OR "DBS" OR "virtual reality therapy" OR "avatar therapy" OR "digital intervention" OR "mHealth" OR "eHealth" OR "psychedelic-assisted psychotherapy" OR "psilocybin") |
| **Psycinfo** | ("auditory hallucination*" OR "hearing voices" OR "voice-hearing" OR "hallucinations, auditory" OR ("voices" AND "psychosis"))<br>AND<br>("schizophrenia" OR schizophren* OR "psychotic disorders" OR "psychosis" OR "schizoaffective |
|  | disorder")<br>AND<br>("treatment outcome" OR "treatment efficacy" OR "therapy" OR "antipsychotic agents" OR "Clozapine" OR "Haloperidol" OR "Olanzapine" OR "Risperidone" OR "Aripiprazole" OR "Cognitive Behavioral Therapy" OR "CBT" OR "cognitive remediation therapy" OR "psychoeducation" OR "family therapy" OR "supportive therapy" OR "electroconvulsive therapy" OR "rTMS" OR "transcranial magnetic stimulation" OR "tDCS" OR "transcranial direct current stimulation" OR "Deep Brain Stimulation" OR "DBS" OR "virtual reality therapy" OR "avatar therapy" OR "digital intervention" OR "mHealth" OR "eHealth" OR "psychedelic-assisted psychotherapy" OR "psilocybin") |
| **Medline** | exp "Auditory Hallucinations" / OR "auditory hallucination".tw OR "hearing voices" / OR "voice-hearing".tw OR ("voices" AND "psychosis").tw<br>AND<br>(exp "Schizophrenia" / OR schizophren*.tw OR exp "Psychotic Disorders" / OR "psychosis" / OR "schizoaffective disorder".tw)<br>AND<br>(exp "Treatment Outcome" / OR "treatment efficacy".tw OR "therapy".tw OR exp "Antipsychotic Agents" / OR "Clozapine".tw OR "Haloperidol".tw OR "Olanzapine".tw OR "Risperidone".tw OR "Aripiprazole".tw OR "Cognitive Behavioral Therapy".tw OR "CBT".tw OR "cognitive remediation therapy".tw OR "psychoeducation".tw OR "family therapy".tw OR "supportive therapy".tw OR exp "Electroconvulsive Therapy" / OR "rTMS".tw OR "transcranial magnetic stimulation".tw OR "tDCS".tw OR "transcranial direct current stimulation".tw OR "Deep Brain Stimulation".tw OR "DBS".tw OR "virtual reality therapy".tw OR "avatar therapy".tw OR "digital intervention".tw OR "mHealth".tw OR "eHealth".tw OR "psychedelic-assisted psychotherapy".tw OR "psilocybin".tw) |
| **Web Of Science** | ("auditory hallucination*" OR "hearing voices" OR "voice-hearing" OR "hallucinations, auditory" OR ("voices" AND "psychosis"))<br>AND<br>("schizophrenia" OR schizophren* OR "psychotic disorders" OR "psychosis" OR "schizoaffective |
|  | disorder") |
|  | AND |
|  | ("treatment outcome" OR "treatment efficacy" OR "therapy" OR "antipsychotic agents" OR "Clozapine" OR "Haloperidol" OR "Olanzapine" OR "Risperidone" OR "Aripiprazole" OR "Cognitive Behavioral Therapy" OR "CBT" OR "cognitive remediation therapy" OR "psychoeducation" OR "family therapy" OR "supportive therapy" OR "electroconvulsive therapy" OR "rTMS" OR "transcranial magnetic stimulation" OR "tDCS" OR "transcranial direct current stimulation" OR "Deep Brain Stimulation" OR "DBS" OR "virtual reality therapy" OR "avatar therapy" OR "digital intervention" OR "mHealth" OR "eHealth" OR "psychedelic-assisted psychotherapy" OR "psilocybin") |

**Fig. S1.**
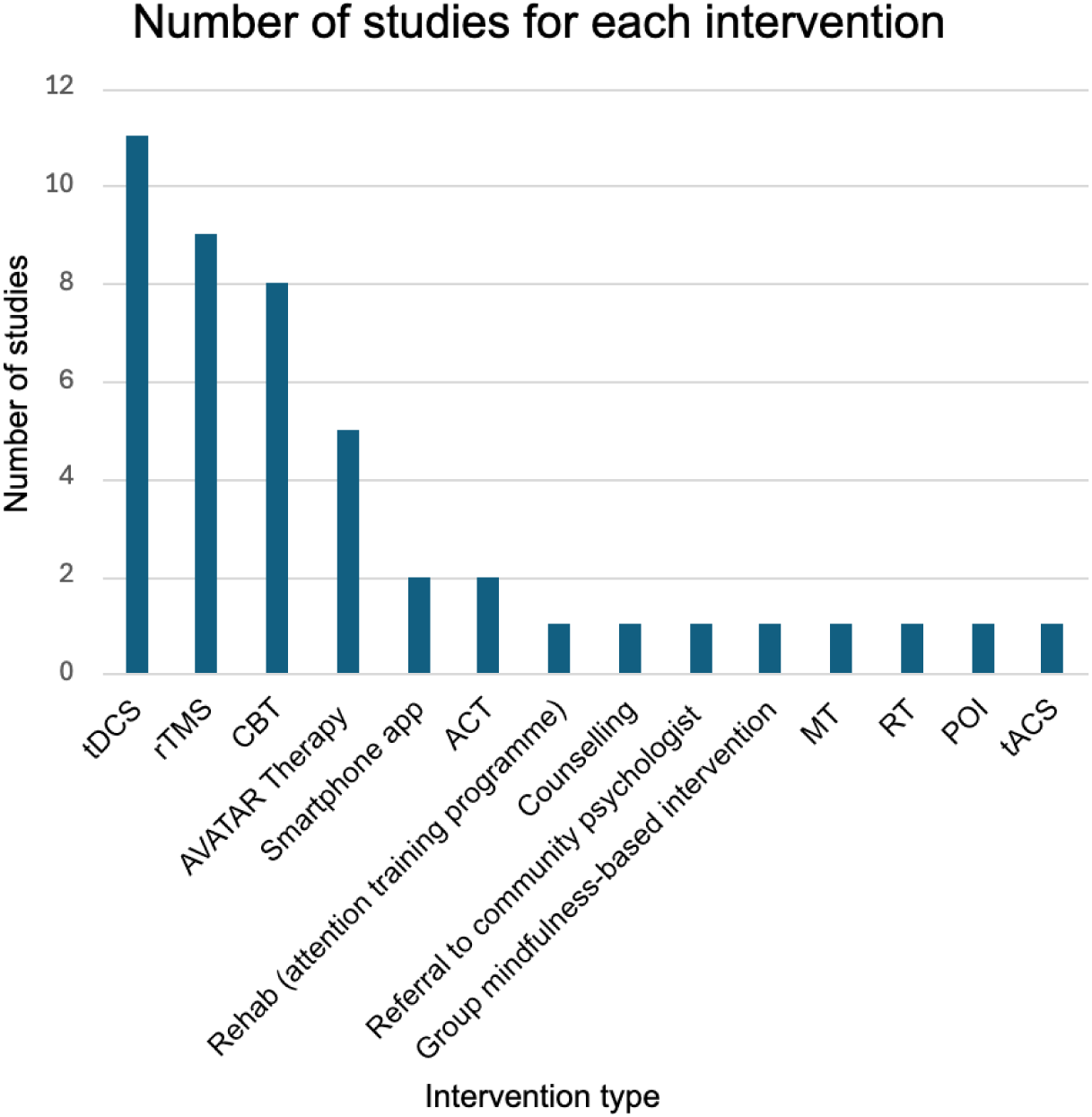
Quantity of each treatment type in the meta-analysis.

**Fig. S2.**
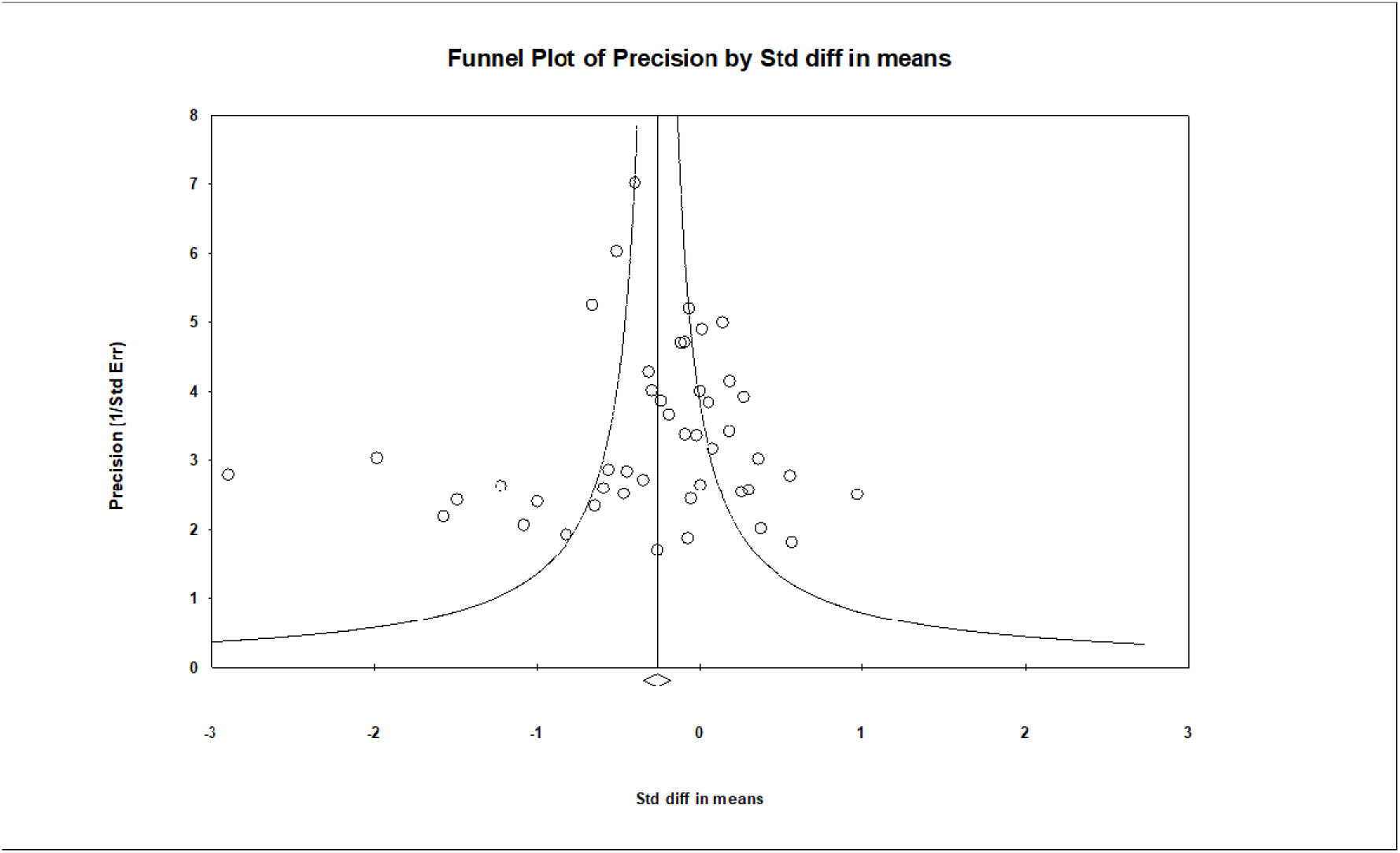
Funnel plot of precision for publication bias.

**Table S3.**
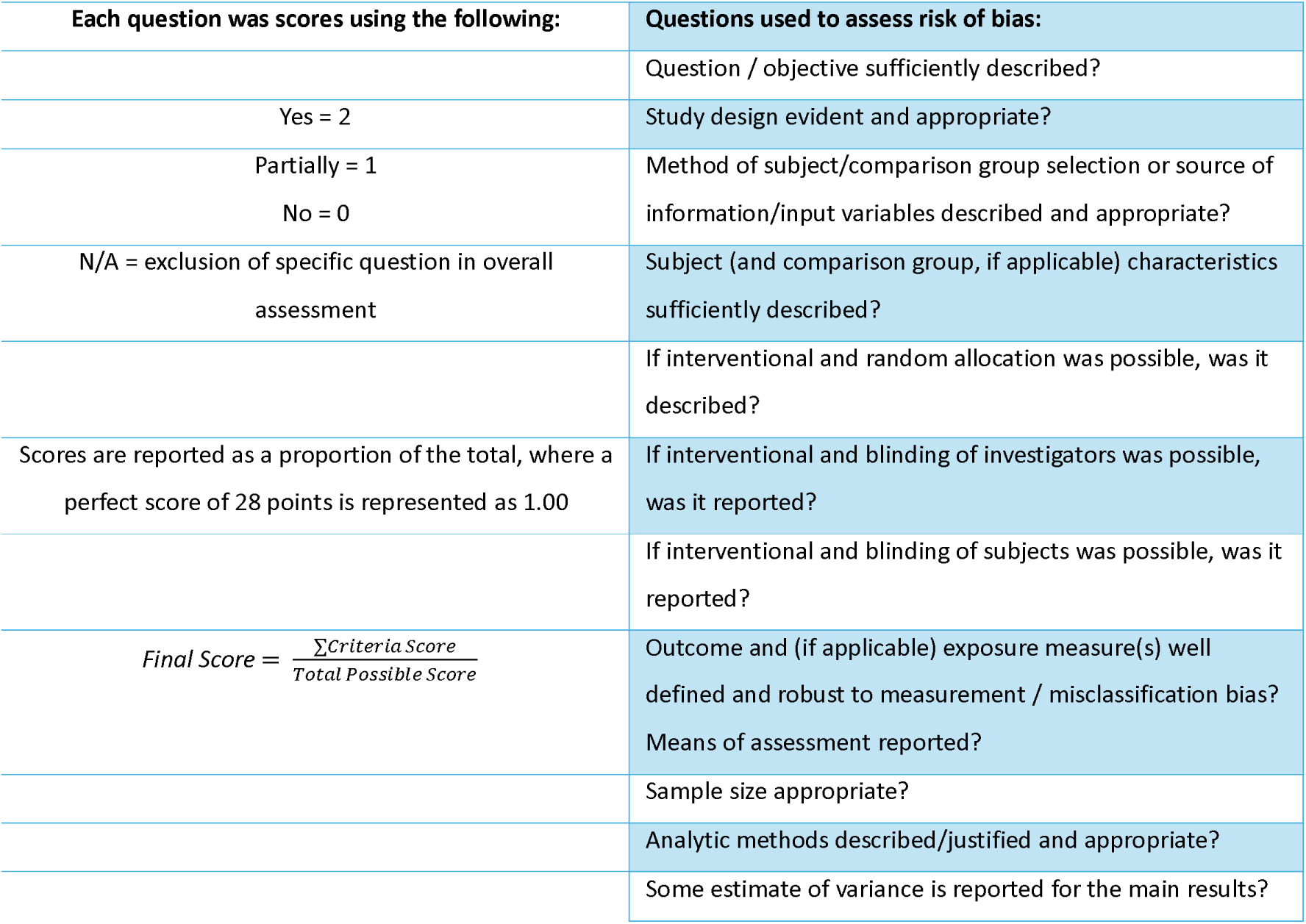
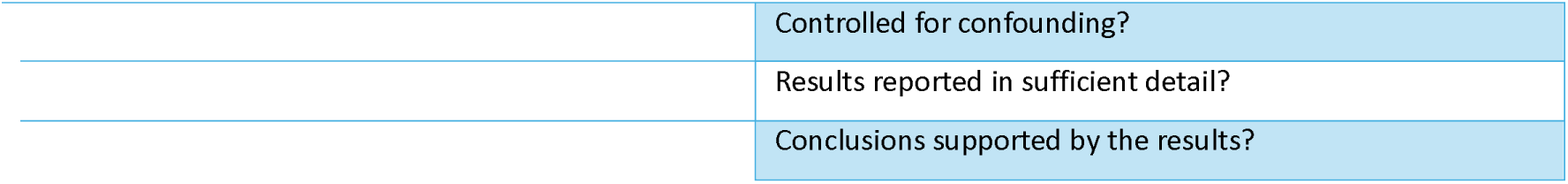
QualSyst risk of bias assessment process.

**Table S4.** QualSyst bias scores for each study.

| Study | QualSyst Bias Score |
| --- | --- |
| Klirova M. et al. 2013 | 0.93 |
| Kråkvik B. et al. 2013 | 0.82 |
| Bais L. et al. 2014 | 0.93 |
| Fitzgerald P. et al. 2014 | 0.93 |
| Freeman D. et al. 2015 | 0.93 |
| Naeem F. et al. 2015 | 0.83 |
| Ray P. et al. 2015 | 0.93 |
| Fröhlich F. et al. 2016 | 0.93 |
| Kimura H. et al. 2016 | 0.86 |
| Koops S. et al. 2016 | 0.93 |
| López-Luengo B. et al. 2016 | 0.86 |
| Mondino M. et al. 2016 | 0.91 |
| Naeem F. et al. 2016 | 0.89 |
| Chadwick P. et al. 2016 | 0.89 |
| Gottlieb J. et al. 2017 | 0.83 |
| Hayward M. et al. 2017 | 0.86 |
| Husain M. et al. 2017 | 0.82 |
| Paillere-Martinot M. et al. 2017 | 0.93 |
| Schnackenberg J. et al. 2017 | 0.72 |
| Shawyer F. et al. 2017 | 0.96 |
| Bose A. et al. 2018 | 0.96 |
| Chang C. et al. 2018 | 0.93 |
| Craig T. et al. 2018 | 0.89 |
| Hazell C. et al. 2018 | 0.82 |
| Koops S. et al. 2018 | 0.93 |
| Mellin J. et al. 2018 | 0.93 |
| Percie du Sert O. et al. 201852 | 0.86 |
| Pinar S. et al. 2018 | 0.70 |
| Kantrowitz J. et al. 201954 | 0.93 |
| Lindenmayer J. et al. 201955 | 0.93 |
| Bell I. et al. 2020 | 0.86 |
| Lüdtke T. et al. 2020 | 0.79 |
| Sevi O. et al. 2020 | 0.86 |
| Valiengo L. et al.2020 | 0.96 |
| Dellazizzo L. et al. 2021 | 0.82 |
| El Ashry A. et al. 2021 | 0.76 |
| Klein H. et al. 2021 | 0.89 |
| Liang N. et al. 2022 | 0.86 |
| Solar A. et al. 2022 | 0.93 |
| Tyagi P. et al. 2022 | 0.93 |
| Zhang M. et al. 2022 | 0.93 |
| Xie Y. et al. 2023 | 0.89 |
| Garety P. et al. 2024 | 0.86 |
| Hua Q. et al. 2024 | 0.96 |
| Jongeneel A. et al 2024 | 0.89 |

